# Locked to the match: subthalamic engagement in sport match viewing

**DOI:** 10.64898/2026.05.20.26353675

**Authors:** Salvatore Falciglia, Laura Caffi, Fabrizio Luiso, Chiara Palmisano, Alberto Mazzoni, Ioannis Ugo Isaias

## Abstract

Adaptive deep brain stimulation (aDBS) of the subthalamic nucleus (STN) ameliorates motor symptoms in advanced Parkinson’s disease (PD) by modulating stimulation in real time using neural signals linked to motor symptoms. Whether these signals also reflect ongoing behavior in naturalistic settings remains unknown. We recorded bilateral STN local field potentials from eight PD patients undergoing aDBS during live-streamed sports viewing and show that low-frequency dynamics encode behavioral engagement across multiple timescales. Engaged viewing increased activity in the stimulation-targeted frequency band relative to control conditions. At a finer timescale, moment-to-moment engagement modulated rapid fluctuations in left STN activity with amplitude maxima and minima time-locked to salient in-match events. These findings reveal that STN activity dynamically reflects real-world behavioral states and establish a foundation for behaviorally informed neuromodulation strategies.

---

Parkinson’s disease (PD) is the second most common neurodegenerative disorder after Alzheimer’s disease, with prevalence increasing at a rate that exceeds demographic aging alone[1]. The disease primarily affects basal ganglia circuits where progressive dopamine loss leads to characteristic motor symptoms, most notably bradykinesia and rigidity[2]. PD also encompasses a broad spectrum of neuropsychiatric symptoms that are increasingly recognized as central contributors to disease burden[3], yet lack well-defined neural correlates for targeted therapeutic interventions. More than one third of patients with PD experience apathy[4] (i.e., diminished goal-directed behavior and emotional responsivity), depression[5, 6], and anhedonia[7] (i.e., inability to experience pleasure or anticipate reward). These symptoms directly compromise the capacity for sustained engagement during emotionally salient experiences[8].

Deep brain stimulation (DBS) of the subthalamic nucleus (STN) represents a mainstay therapeutic intervention for advanced PD with medication-refractory motor fluctuations[9]. These fluctuations include transitions between disabling “off” and “on” states, as well as levodopa-related dyskinesias, that substantially impair quality of life. Conventional DBS (cDBS) delivers continuous stimulation with fixed, predefined parameters[10]. Next-generation implantable pulse generators (IPGs) enable the simultaneous recording of neural activity[11], transforming DBS into a bidirectional neural interface and positioning it within the emerging class of implantable neuroprosthetic systems. This capability creates unprecedented opportunities for adaptive DBS (aDBS), in which stimulation is dynamically adjusted in real time based on neural biomarkers sensed directly from implanted electrodes[12].

Neural biomarkers currently used for aDBS in PD are local field potentials (LFPs) recorded directly from implanted DBS electrodes. In particular, STN-LFPs oscillatory activity in the beta band (12-30 Hz), defined within a patient-specific frequency range, has emerged as a robust correlate of motor impairment, especially bradykinesia and rigidity[13]. Beta power increases with symptom severity and decreases with clinical improvement following both levodopa administration and STN DBS[14, 15].

Recent clinical studies have shown that patients eligible for aDBS prefer this modality over cDBS[16, 17], reporting improvements extending beyond motor symptoms to non-motor domains and overall well-being. The STN constitutes a key integrative node within basal ganglia circuitry, supporting motor, limbic, and cognitive processing[18]. Consistently, the effects of STN DBS extend beyond motor circuits to influence non-motor domains, including affective and cognitive functions, reflecting a neuromodulatory impact that is inherently multimodal and distributed across large-scale brain networks. Direct evidence for this broader impact is provided by acute STN DBS-related adverse effects, including alterations in mood, impaired emotional recognition and expression, impulse control disturbances, and cognitive changes. To date, these aspects have been investigated primarily in controlled laboratory settings[19]. Such studies demonstrate that emotional information across sensory modalities (e.g., visual and auditory) is encoded in STN spectral dynamics across multiple frequency bands[20, 21], and that distinct affective dimensions, such as valence and arousal, are dissociated at both population and single-neuron levels[22–24]. Long term studies on STN dynamics during daily activities in home environments[25–28] have mainly focused on motor fluctuations[29], sleep-wake transitions, and circadian rhythms[30], whereas emotional modulation has remained largely unexplored.

The gap between laboratory characterization and naturalistic behavior represents a critical challenge for both the mechanistic understanding of STN function and the development of next-generation neuromodulation strategies. aDBS algorithms should not be limited to adaptating stimulation to specific symptoms, but rather should operate within the full complexity of real-world behavior, capturing not only motoractivity but also social, cognitive, and limbic processes, as well as their associated symptom fluctuations[31]. For instance, cognitive overload and anxiety are well known to exacerbate tremor and gait disturbances, particularly freezing of gait[29, 32].

For neuromodulation, this translates to a critical requirement: biomarkers must be validated not only during laboratory tasks but throughout sustained, multidimensional experiences that characterize everyday life[33].

Chronic sensing technologies now enable continuous neural monitoring during unconstrained behavior[16], enabling the identification of ecologically grounded neural signatures suitable for tailored aDBS strategies. These advances provide unprecedented opportunities to characterize neural dynamics in naturalistic contexts and to identify signal features that reliably disambiguate behavioral states.

Here, we investigate STN dynamics in chronically aDBS-treated PD patients during the viewing of high-stakes live sporting events. This naturalistic paradigm provided an ecologically valid context in which individuals experience prolonged sequences of anticipation, excitement, disappointment, and satisfaction, reflecting genuine emotional engagement embedded within continuous attentional demands, outcome prediction, working memory for game state, and narrative integration. By leveraging chronic, bilateral STN-LFP recordings in real-world conditions, we directly probe how motor and affective processes co-modulate control-relevant neural signals, informing the design of next-generation adaptive neuroprosthetic therapies in PD.

## Results

### Ecological monitoring of subthalamic activity during sports viewing

To investigate how engagement modulates basal ganglia activity in real-world conditions, we recorded STN-LFPs in eight patients with PD treated with chronic STN aDBS while they viewed sports matches either as engaged supporters or as neutral viewers (Fig. 1a; Methods; fig. Extended Data 1, Extended Data 2; tables Supplementary Information 1 - Extended Data 2).

**Fig. 1:**
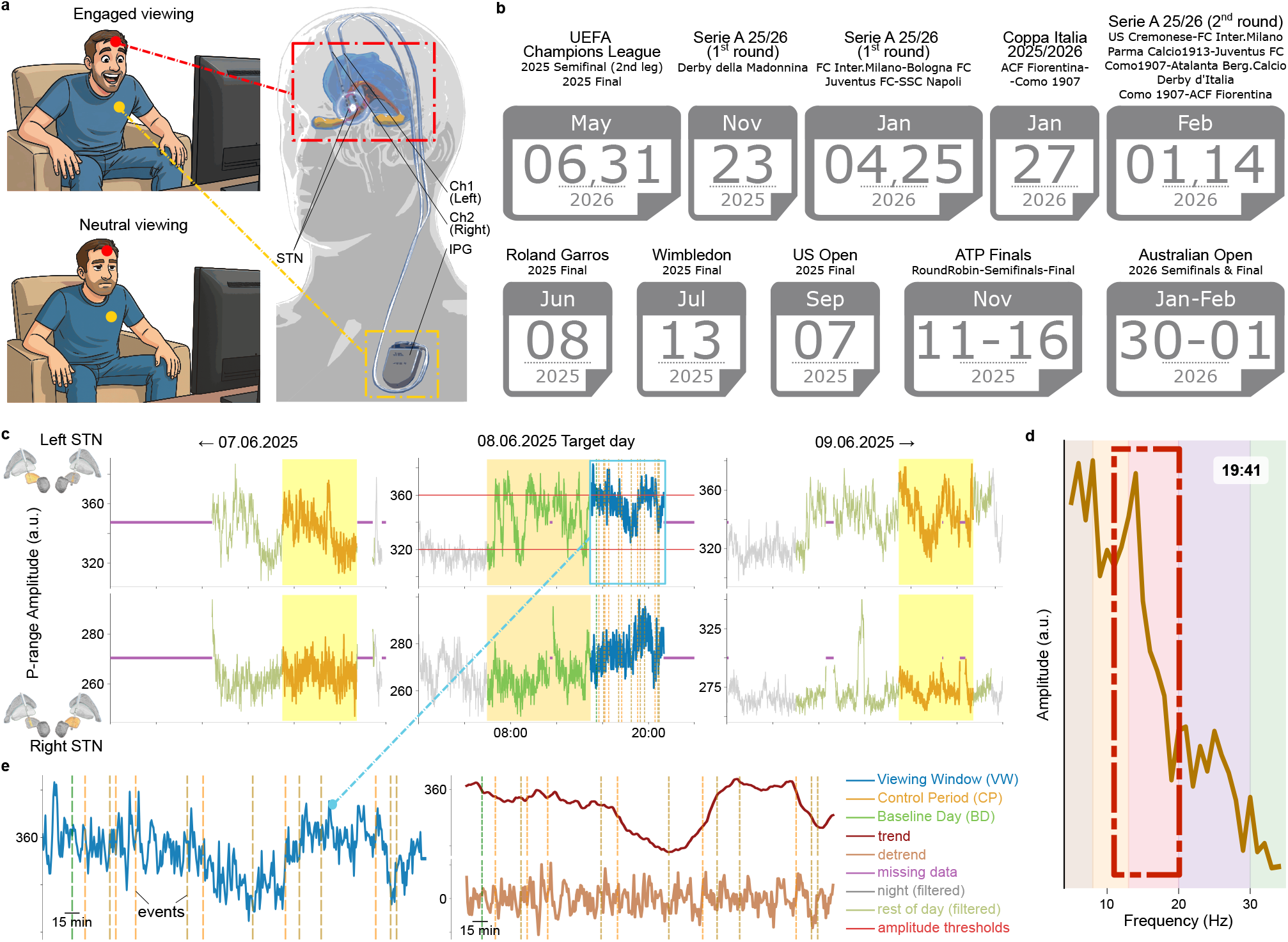
Experimental paradigm and multilevel analysis structure. **a**, Eight patients with Parkinson’s disease underwent subthalamic nucleus (STN) deep brain stimulation (DBS) with an implantable pulse generator (IPG, the AlphaDBS device by Newronika SpA) located on the chest and two leads implanted in the left (Ch1) and right (Ch2) STN, both stimulating and recording (tables Supplementary Information 1 - Extended Data 2; fig. Extended Data 1 for post-surgery lead-group reconstruction). Patients watched one or more live, high-stakes tennis and soccer matches while seated at home in front of the television and asked to minimize motor activity. Based on post-viewing interviews (tables Extended Data 3, Supplementary Information 2), viewing sessions were classified as engaged or neutral. One patient additionally watched five matches at the stadium, where the context differed substantially due to different external conditions and motor activity levels. These viewing sessions were analyzed separately as a test case. **b**, Soccer matches included UEFA Champions League 2025 Semifinal (second leg) and Final, several Serie A 2025/2026 matches, and one Coppa Italia 2025/2026 match. Tennis matches included Roland Garros 2025 Final, Wimbledon 2025 Final, US Open 2025 Final, multiple ATP Finals 2025 matches, and Australian Open 2026 Semifinals and Final. **c**, Patients’ STN local field potential (LFP) amplitude was recorded chronically and bilaterally with 1-min resolution in a personalized frequency band (P-range) comprising part of alpha and low beta frequencies (5-23 Hz). For stimulation, a driver channel was chosen, for which two amplitude thresholds were fixed according to the IPG’s stimulation algorithm. We monitored the target match day together with a variable period of days before and after, excluding nighttime (10pm - 6am) (fig. Extended Data 2). Intervals of missing data in the recordings - with stimulation still on - were present in each day of monitoring, mainly due to device charging. Based on these data, we defined three data categories: the viewing window (VW), P-range amplitude during the match ±5 min on the target day; the control period (CP), P-range amplitude from the same time window on the remaining days of monitoring; the baseline day (BD), P-range amplitude on the target day excluding VW. Data from patient NWKb during the Roland Garros 2025 Final are visualized. **d**, AlphaDBS recordings saved full amplitude spectra (5-34 Hz) with 10-min resolution for each channel, from which the corresponding P-range amplitude value of that minute could be retrieved. For VW, CP, and BD, amplitude time series with 10-min resolution were extracted for the classic frequency bands: theta 5-8 Hz, alpha 8-12 Hz, low-beta 12-20 Hz, high-beta 20-30 Hz, gamma 30-34 Hz. **e**, An additive decomposition model applied to the VW signal at 1-min resolution distinguishes between trend and detrended components, allowing for the study of STN P-range amplitude around in-match events. The trend was estimated using a centered moving average 30-min window. In our working hypothesis, (i) the trend component reflects baseline shifts and long-term neural dynamics (slower timescales), while (ii) the detrended component captures event-specific, transient neural activations (faster timescales).

Patients were monitored during naturalistic viewing in their home environment, seated comfortably while watching live, high-stakes tennis or soccer matches (UEFA Champions League 2025 Semifinal 2nd leg and Final, Roland Garros 2025 Final, Wimbledon 2025 Final, US Open 2025 Final, matches from the ATP Finals 2025, matches from the Serie A 2025/2026 and the Coppa Italia 2025/2026, Australian Open 2026 Semifinals and Final; Fig. 1b, Methods). We chronically recorded bilateral STN activity (Ch1/Left, Ch2/Right) in 55 of 63 monitoring periods and unilaterally in 8/63 (Methods). One patient (NWKc) additionally attended five live soccer matches in stadium settings (UEFA Champions League 2025 Semifinal 2nd leg and Final, Serie A 2025/2026 1st round Derby della Madonnina, Serie A 2025/2026 1st round FC Internazionale Milano vs Bologna FC, and Serie A 2025/2026 2nd round Derby d’Italia) and was analyzed separately given the different motor and sensory conditions compared to home viewing (Fig. 1b; Methods).

Viewing windows (VWs) were defined from 5 min before match onset to 5 min after match end. VWs were retrospectively categorized into two categories based on post-match patient interviews (Fig. 1a; Methods; tables Extended Data 3, Supplementary Information 2): engaged (VWs while rooting for one of the contenders; *n* = 29 viewings across 6 patients) and neutral (VWs without rooting for either contender; *n* = 17 viewings across 8 patients).

Engaged and neutral VWs were each compared with a control period (CP) and a baseline day (BD) activity. CP was defined relative to each VW as the same time interval on the three days before and after the event (Fig. 1c; fig. Extended Data 2; Methods). BD was defined as neural activity on the match day outside the corresponding VW and nighttime hours (10pm-6am) (Fig. 1c; fig. Extended Data 2; Methods). Finally, in a similar fashion, time-matched control intervals from patients not watching the match (n=12 recordings across 6 patients) were considered for comparative analyses (VW-CP and VW-BD; Methods). For consistency, both viewing and concurrent non-viewing windows are hereafter denoted as VW.

For each patient, analysis first focused on a personalized frequency range (P-range) within the [5-23] Hz alpha-beta range (Fig. 1d; table Extended Data 1), defined based on patient-specific clinical criteria and used to modulate aDBS current (Methods). P-range amplitude was estimated at 1-min resolution across the entire monitoring period (Fig. 1c; fig. Extended Data 2). Complementary analyses examined amplitude spectra within the [5, 34] Hz range at 10-min resolution (Fig. 1d; fig. Extended Data 4a, b).

During engaged VWs, we further compared the temporal evolution of STN activity at salient in-match events with that observed during the remainder of the match (Fig. 1e; Methods). This framework enabled the dissociation of engagement-related effects across two timescales: a sustained component reflecting overall engagement during viewing and a transient component capturing rapid neural dynamics associated with salient in-match events.

### Subthalamic P-range amplitude increases during engaged viewing

We first assessed the average effect of engaged match viewing by comparing P-range amplitude during VW with BD and CP within engaged (*n* = 29), neutral (*n* = 17), and non-viewing (*n* = 12) categories.

We performed per-match, per-hemisphere VW-CP and VW-BD comparisons (VW *n* = [52, 388], CP *n* = [164, 3180], BD *n* = [374, 881], where n denotes the [min, max] number of samples within each window type; Mann-Whitney U test, Benjamini-Hochberg FDR correction) and a population-level analysis of paired medians (Wilcoxon signed-rank test, Benjamini-Hochberg FDR correction) (Fig. 2). A two-way ANOVA to test the interaction between viewing and engagement was not feasible because engagement was only defined during viewing (Methods), precluding a full crossing of the two factors; categories were therefore analyzed separately. To account for repeated measures within patients (*n* =6-8 patients, 1-11 viewings each), we implemented linear mixed-effects models, including condition (VW vs. pooled reference BD and CP) as a fixed effect and patient as a random intercept (Methods).

**Fig. 2:**
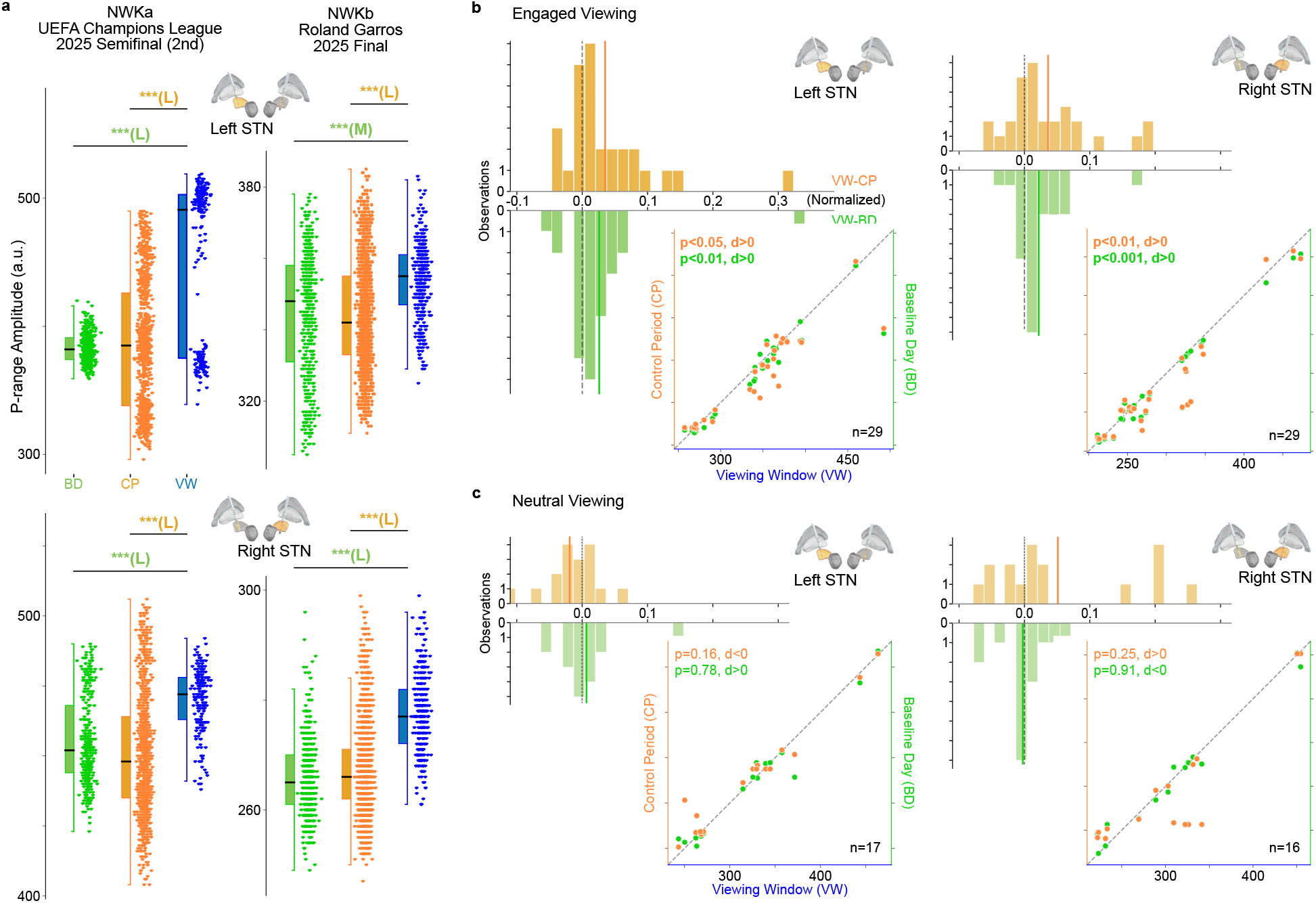
Engagement modulates subthalamic P-range amplitude during real-world sport match viewing. **a**, VW-CP and VW-BD comparisons were investigated to assess the global engagement effect of the high-stakes sport matches. Per-match, per-channel differences were tested over all P-range amplitude values using the Mann-Whitney U test with Benjamini-Hochberg FDR correction. Cohen’s d quantified effect size, distinguishing negligible (|*d*| < 0.1), small (S, |*d*| ≥ 0.1), medium (M, |*d*| ≥ 0.3), or large (L, |*d*| ≥ 0.7) magnitude of the observed effect. Comparisons for the engaged VWs resulted in 18/29 sessions showing significant (*p* < 0.001) positive medium/large effects in both hemispheres against at least one control condition, indicating increased STN P-range amplitude while watching live sports. Results for patients NWKa and NWKb while watching the UEFA Champions League 2025 Semifinal (2nd leg) and the Roland Garros 2025 Final respectively are shown. Results for all VWs across engagement levels are reported in table Supplementary Information 3. **b, c**, Wilcoxon signed-rank tests (paired, non-parametric) were applied to median pairs for VW-CP and VW-BD for the two categories of engaged VWs (n=29 viewings, bilateral, 6 patients) and neutral VWs (n=17 viewings, bilateral, 8 patients). Significant positive effects (*p* < 0.05) were observed in both channels for both controls only within the engaged VWs category, while no significant effects were found in the neutral VWs category. Population-level analysis results were confirmed through linear mixed effects modeling. Results are shown in two formats for both hemispheres: (i) scatter plot with diagonal line of equality, with two y-axes for CP and BD P-range amplitude values and one x-axis for VW P-range amplitude values (points below the diagonal show higher VW values compared to the associated control); (ii) histogram with null line of equality showing the normalized difference between VW and the associated control P-range amplitude values (bars at positive values of the normalized differences show occurrences of sessions presenting higher VW values compared to the associated control). Results for the engagement effect across viewing and non-viewing categories are shown in fig. Extended Data 3. Results for the engagement effect across frequency bands are shown in fig. Extended Data 4.

Engaged VWs resulted in 18/29 sessions showing significant (*p* < 0.001) positive medium-to-large effect (*d* ≥ 0.3) in both hemispheres (Fig. 2a) for at least one between VW-CP and VW-BD and 4/29 sessions showing significant effect in only one hemisphere. Small-to-negligible effect (*d* < 0.3) was observed in 5/29 sessions for both VW-CP and VW-BD. Significant negative effect in both hemispheres occurred in 2/29 sessions for at least one between VW-CP and VW-BD (table Supplementary Information 3).

Comparison between neutral VWs and CP and neutral VWs and BD yielded non-significant (*p* > 0.05) or small-to-negligible effects, with medium-to-large decreases inconsistent across hemispheres (table Supplementary Information 3). Consistently, the non-viewing category showed no significant deviations in neural activity for both VW-CP and VW-BD (table Supplementary Information 3).

Population-level analysis confirmed these findings as engagement-specific effect, showing significant increase in P-range amplitude associated with engaged viewing in both hemispheres both with respect to CP (*n* = 29; left: *p* < 0.05, *d* = 0.45; right: *p* < 0.01, *d* = 0.45) and BD (*n* = 29; left: *p* < 0.01, *d* = 0.48; right: *p* < 0.001, *d* = 0.69) (Fig. 2b). No significant effects were observed either in the neutral ([VW-CP] left: *n* = 17, *p* = 0.16, *d* < 0; right: *n* = 16, *p* = 0.25, *d* > 0 - [VW-BD] left: *n* = 17, *p* = 0.78, *d* > 0; right: *n* = 16, *p* = 0.91, *d* < 0) (Fig. 2c) or the non-viewing categories ([VW-CP] left: *n* = 12, *p* = 0.79, *d* > 0; right: *n* = 7, *p* = 0.75, *d* < 0 - [VW-BD] left: *n* = 12, *p* = 0.82, *d* < 0; right: *n* = 7, *p* = 0.47, *d* > 0) (fig. Extended Data 3).

Significant results were confirmed through linear mixed effects modelling (model coefficient *β*, Methods) to evaluate intra-patient contribution to the observed effect. The left STN showed significant elevated P-range amplitude during VWs compared to pooled reference (*β* = 9.53 a.u., 95% CI [0.20, 18.85], *z* = 2.00, *p* < 0.05). The right STN showed a similar, non-significant trend (*β* = 10.28 a.u., 95% CI [-1.66, 22.22], *z* = 1.69, *p* = 0.09), with a medium effect size (Cohen’s *d* = 0.39). Intraclass correlation coefficients were high in both hemispheres (left ICC = 86.8%, right ICC = 89.6%; Methods). This indicates that P-range amplitude variance primarily reflected stable, inter-individual differences in chronic STN activity. Consequently, within-patient variation across engaged and neutral viewing conditions accounted for only 11-14% of the total variance (Discussion). These results indicate increased STN-LFPs P-range amplitude during engaged viewing at home of sport matches.

Population-level analysis was also performed on STN-LFPs amplitude within standard frequency bands (theta: (5, 8) Hz, alpha: (8, 12) Hz; low-beta: (12, 20) Hz; high-beta: (20, 30) Hz; gamma: (30, 34) Hz) using 10-min resolution amplitude spectra. Low beta was the only band showing significant increased amplitude after FDR correction, in both VW-CP (right: *p* < 0.05, *d* = 0.31; only a trend in the left STN: *p* = 0.14, *d* = 0.17) and VW-BD comparisons (right: *p* < 0.01, *d* = 0.31; only a trend in the left STN: *p* = 0.05, *d* = 0.38) (fig. Extended Data 4).

Because all patients received aDBS therapy (Methods), we examined whether the increase in P-range amplitude during engaged viewing was accompanied by changes in the stimulation (fig. Extended Data 7). At the population level, bilateral stimulation current (1-min resolution) followed a pattern partially consistent with P-range amplitude fluctuations. Engaged VW was associated with significantly higher stimulation than BD (left: *p* < 0.01, *d* > 0, mean difference= 0.086 mA; right: *p* < 0.05, *d* > 0, mean difference= 0.083 mA; Wilcoxon signed-rank test, *n* = 29), whereas comparisons with CP showed only trends of significance (left: *p* = 0.07, *d* > 0, mean difference= 0.021 mA; right: *p* = 0.11, *d* > 0, mean difference= 0.024 mA). The increase in VW compared to BD (~ 0.09 mA) was modest but consistent across hemispheres (Discussion).

Comparative analysis showed no difference in stimulation current either in the neutral ([VW-BD] left: *p* = 0.58, *d* < 0, mean difference= −0.024 mA; right: *p* = 0.86, *d* < 0, mean difference= −0.006 mA - [VW-CP] left: *p* = 0.32, *d* > 0, mean difference= 0.006 mA; right: *p* = 0.32, *d* > 0, mean difference= 0.006 mA; Wilcoxon signed-rank test, *n* = 17) or the non-viewing categories ([VW-BD] left: *p* = 0.94, *d* > 0, mean difference= 0.017 mA; right: *p* = 0.93, *d* > 0, mean difference= 0.021 mA - [VW-CP] left: *p* = 0.5, *d* > 0, mean difference= 0.042 mA; right: *p* = 0.34, *d* > 0, mean difference= 0.033 mA; Wilcoxon signed-rank test, *n* = 12).

### Left subthalamic P-range amplitude and in-match events show temporal coupling

Soccer matches lasted 95-100 minutes, while tennis matches lasted much longer (1h30min - 5h29min). We investigated whether changes in the P-range amplitude dynamics in the 1-min timescale temporally aligned with in-match particularly engaging phases/events (Methods). Both kinds of match are highly inhomogeneous in terms of density of subjective and or objective key moments (Methods).

Twenty-seven engaged VW from 6 patients (Methods) were partitioned in higher (HE) versus lower (LE) engagement intervals based on post-match interviews (Fig. 3a; table Supplementary Information 2). On a finer temporal scale, key-event times within each live match were retrieved with 1-min resolution (Fig. 1e; Fig. 3a; Methods).

**Fig. 3:**
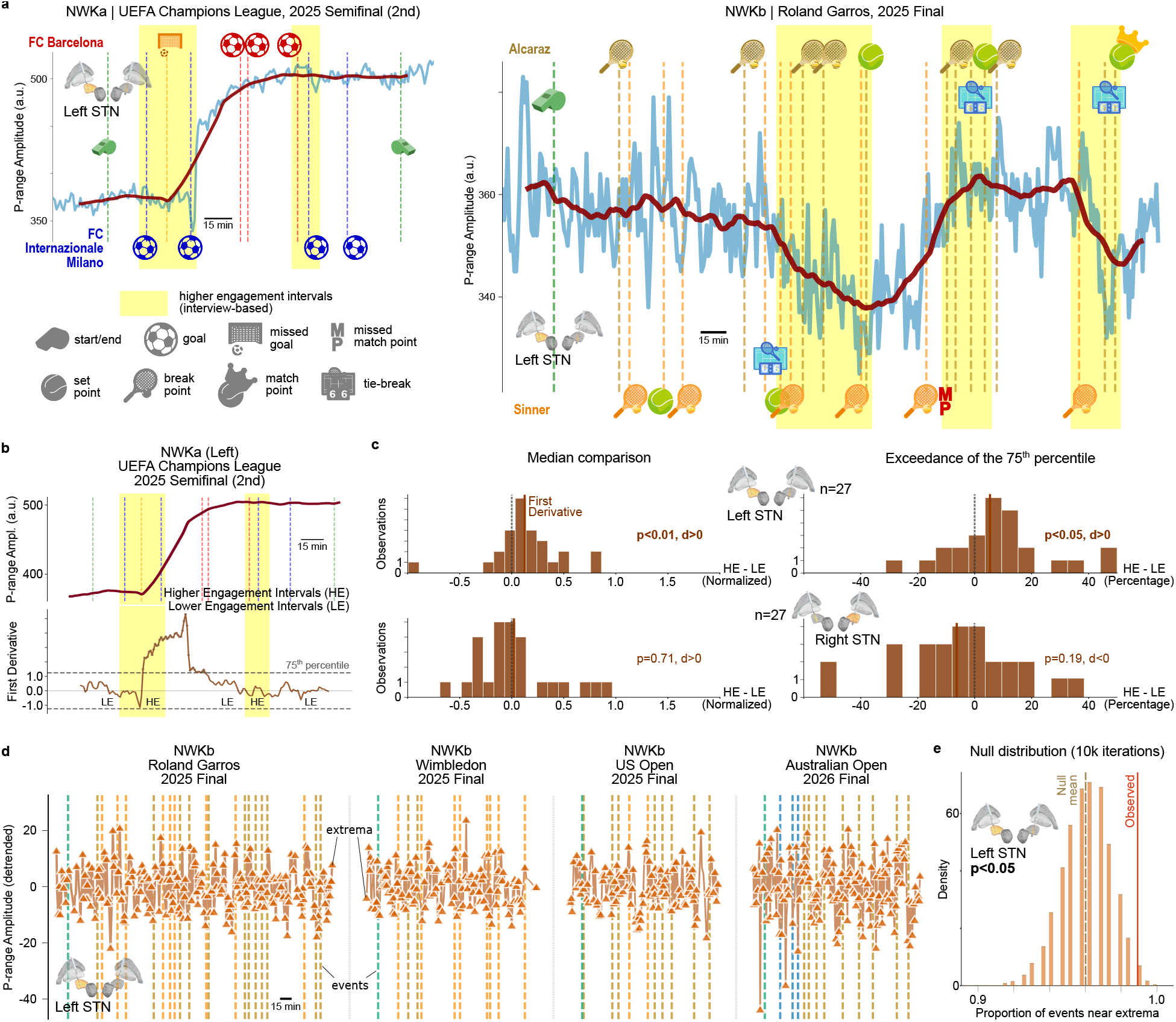
Subthalamic P-range amplitude tracks in-match engagement during home viewing. **a**, STN P-range amplitude trend profiles of engaged VWs are computed using a centered moving average 30-min window, showing the slow changes in the P-range amplitude in correspondence of in-match events. Higher engagement intervals were identified within engaged VWs according to the recalled critical match moments during post-viewing interview (tables Extended Data 3, Supplementary Information 2). Data from left STN of patients NWKa and NWKb are visualized while watching the UEFA Champions League 2025 Semifinal (2nd leg) and the Roland Garros 2025 Final respectively. **b**, Specifically, engaged VWs were further partitioned in higher (HE) and lower (LE) engagement intervals based on post-viewing interviews. Trend’s derivatives were estimated through the Savitzky-Golay Filter and evaluated through pairs of median values (HE, LE) extracted for each VW computing their absolute-valued magnitude and the percentage of time points exceeding their 75^*th*^ percentile threshold. First derivative of the left STN P-range trend from patient NWKa while watching UEFA Champions League 2025 Semifinal (2nd leg) is visualized (second derivative in fig. Extended Data 5). **c**, Wilcoxon signed-rank tests (paired, non-parametric) were applied to median pairs for HE-LE for the category of engaged VWs (n=29). Significant positive effects (*p* < 0.05) were observed only in the left hemispheres, evaluating both absolute-valued magnitude and percentage of time points exceeding threshold. Population-level analysis results are shown for first derivative (fig. Extended Data 5 for second derivative) in the histogram format with null-line of equality showing the normalized difference HE-LE for absolute-valued magnitude and the difference in percentage for the threshold exceedance. **d**, STN P-range amplitude detrended components allow for studies over coupling between the neural signal and the in-match events. Multiple VWs within individual patients were aggregated to increase statistical power, under the hypothesis of similar within-patient engagement for all engaged VWs recorded. Extrema detection identified local maxima and minima with minimum height threshold of 0.5 relative to neighboring points. Data from left STN of patient NWKb while watching the four Slam finals and in-match events are visualized. **e**, Non-parametric permutation testing with 10K iterations allowed to generate null distributions for both hemispheres evaluating extrema-near-events proportion and maximum variation-at-events. Statistical significance was assessed through one-tailed (right-tailed) tests and reached only with the extrema-near-events proportion metric on the left STN.

To assess whether HE was associated to faster variations in P-range amplitude, we computed the first time derivative of the slow trend component of the 1-min resolution P-range amplitude in VW (Fig. 3b). Population-level analysis of the absolute-valued median trend’s time derivatives (Wilcoxon signed-rank test and Linear Mixed Effects modelling; Methods) revealed that HE intervals exhibited steeper P-range amplitude fluctuations than LE intervals, a result restricted to the left hemisphere (left: *p* < 0.01, *d* = 0.44; right: *p* = 0.71, *d* = 0.05). Similarly, the time spent by the signal time derivative exceeding the 75^*th*^ percentile was significantly higher during HE only in the left STN (left: 30±16% vs 25±11%, *p* < 0.05; right: 23±15% vs 28±11%, *p* = 0.19) (Fig. 3c). The second time derivative of the slow trend component was also computed (fig. Extended Data 5a), yielding consistent results at the population level (fig. Extended Data 5b).

Linear mixed effects modelling (model coefficient *β*, Methods) further confirmed our results, showing only in the left STN a significant more elevated absolute-valued first trend time derivative in HE intervals (left: *β* = 0.1 a.u., 95% CI [0.013, 0.188], *z* = 2.25, *p* < 0.05, ICC = 2.9%; right: *β* = −0.002 a.u., 95% CI [-0.090, 0.087], *z* = −0.04, *p* = 0.97, ICC = 46.6%). Similar results held for the left STN absolute-valued second time derivative (left: *β* = 0.040 a.u., 95% CI [0.009, 0.069], *z* = 2.57, *p* < 0.05, ICC = 0.5%; right: *β* = 0.004 a.u., 95% CI [-0.025, 0.033], *z* = 0.25, *p* = 0.80, ICC = 37.4%) and proportion of time points exceeding the 75^*th*^ percentile (left: *β* = 7.97 percentage points, 95% CI [2.29, 13.65], *z* = 2.75, *p* < 0.01, ICC = 10.4%; right: *β* = 2.35 percentage points, 95% CI [-1.92, 6.62], *z* = 1.08, *p* = 0.28, ICC = 12.3%).

Notably, P-range amplitudes were highly heterogeneous between patients (ICC = 86.8-89.6%), yet their temporal dynamics (i.e., time derivatives) were remarkably homogeneous (ICC = 0.5-2.9%). This suggests that the rate and acceleration of P-range amplitude changes are consistent across patients despite large differences in P-range amplitude levels (Discussion).

For the three patients for which we collected a sufficiently high number of events (NWKb *n* = 12; NWKc *n* = 8 with five of them at the stadium; NWKd *n* = 5; Methods) we investigated the relationship between instantaneous events and signal extrema (local maxima/minima) in the fast detrended component (Fig. 3d; Fig. 1e; Methods). Extrema-near-events metric revealed significant association between time of key-events and left P-range amplitude maxima or minima (permutation testing, 10K iterations; NWKb 186 events; left: *n* = 1647 extrema, *p* < 0.05, *d* = 2.12; right: *n* = 1419 extrema, *p* = 0.59, *d* = −0.06 – NWKc 130 events; left: *n* = 898 extrema, *p* < 0.05, *d* = 1.77; right: *n* = 710 extrema, *p* = 0.75, *d* = −0.40 – NWKd 58 events; left: *n* = 473 extrema, *p* < 0.05, *d* = 1.97; right: *n* = 439 extrema, *p* = 0.56, *d* = 0.02) (Fig. 3e). Maximum variation-at-events metric revealed significant P-range amplitude excursions at event times only for one out of these three patients in the right STN (permutation testing, 10K iterations; NWKc right: *p* < 0.005, *d* = 2.73; Methods).

These results suggest STN P-range amplitude in the left hemisphere is modulated on the minutes timescale by particularly engaging moments/events.

### Subthalamic correlates of engagement during in-stadium sport viewing

The *n* = 5 engaged VWs recorded from one patient at the stadium (NWKc; Fig. 4a; chronological order: UEFA Champions League 2025 Semifinal and Final, Serie A 2025/2026 Derby della Madonnina, FC Inter.M.-Bologna FC, and Derby d’Italia) were analyzed descriptively given the limited number of viewings, except for event-related metrics where permutation testing was applied. These VWs did not exhibit consistently the higher P-range amplitude observed at-home during engaged viewing (VW-CP: left −0.04, 0.06, −0.08, −0.05, −0.08 a.u., right −, −, 0.01, −0.01, −0.02 a.u.; VW-BD: left 0.00, −0.01, 0.00, 0.01, −0.04 a.u., right −, −, 0.05, 0.03, −0.02 a.u.; Fig. 4b). However, the relationship between STN activity and engagement levels on faster timescale was coherent with what observed at-home during engaged viewing. In the left STN, trend time derivatives were higher during HE than during LE intervals (first: 0.18, 0.14, 0.07, 0.31, 0.17 a.u.; second: 0.13, 0.14, 0.09, 0.17, −0.09 a.u.; Fig. 4c) as well for the proportion of time points exceeding the 75^*th*^ percentile (first: 16.7, 8.6, 9.0, 35.7, 16.8 percentage points; second: 8.8, 19.0, 15.8, 13.6, −18.5 percentage points; Fig. 4c). Extrema-near-events metric revealed a trend of association between key events and extrema in the detrended components (permutation testing, 10K iterations; *n* = 5 VWs, *p* = 0.07, *d* = 1.61). In addition, maximum variation-at-events metric revealed a trend of higher amplitude excursions at event times in the right STN (permutation testing, 10K iterations; *n* = 3 VWs, *p* = 0.13, *d* = 1.11; Methods). This suggests that the engagement-related P-range amplitude changes might depend on the context, but within match fluctuations might be largely independent from it.

**Fig. 4:**
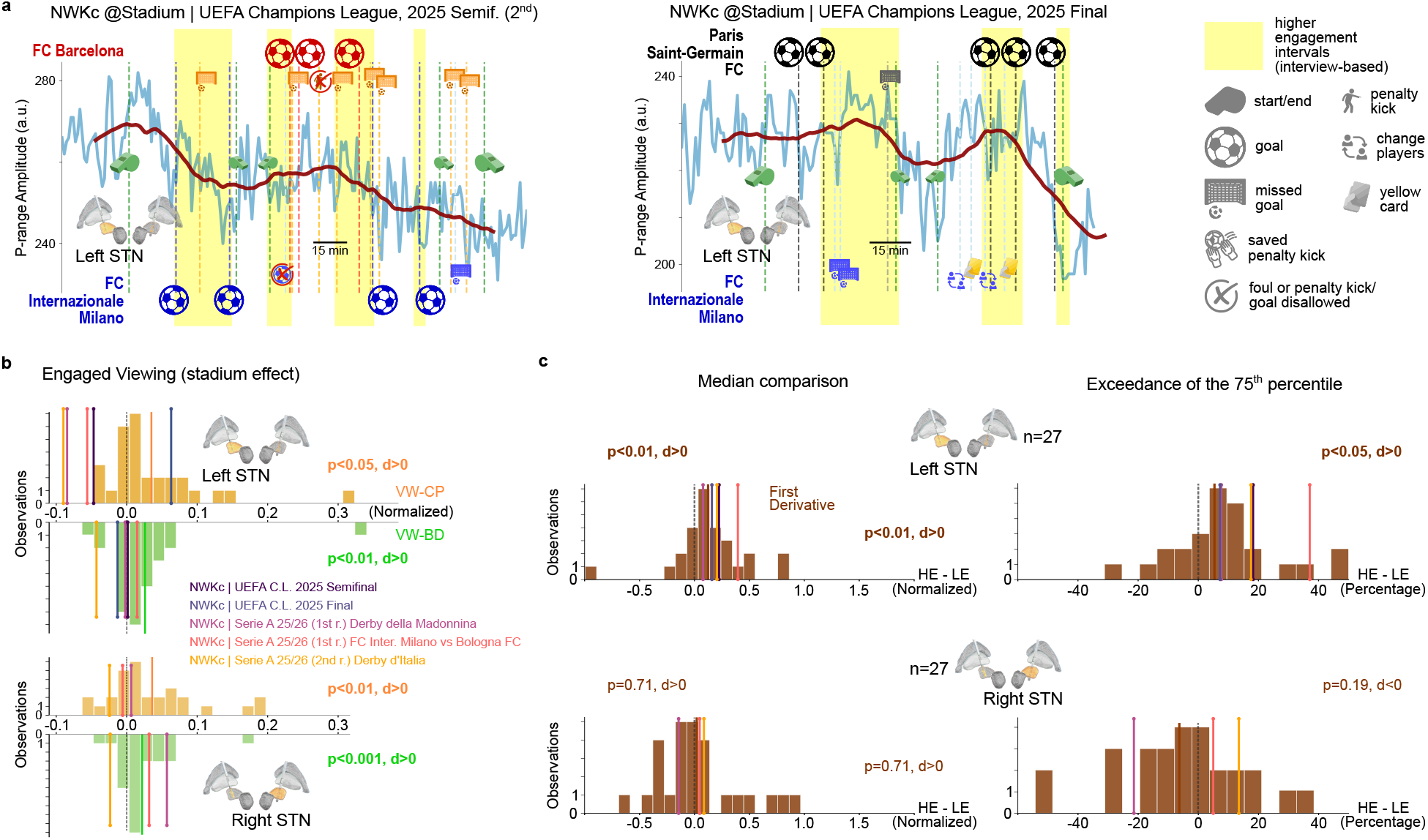
Subthalamic P-range amplitude tracks in-match engagement during stadium viewing. **a**, STN P-range amplitude and trend profiles of the five engaged VWs (chronological order: UEFA Champions League 2025 Semifinal and Final, Serie A 2025/2026 Derby della Madonnina and FC Inter.M.-Bologna FC) of patient NWKc at the stadium with higher engagement intervals highlighted based on the post-viewing interview. Data from the left STN are visualized. **b**, Normalized difference of VW and the associated control P-range amplitude values for the five stadium VWs contextualized within the engaged home viewing population results. Right STN recordings were available only for three VWs (Serie A 2025/2026 Derby della Madonnina, FC Inter.M.-Bologna FC, and Derby d’Italia). Results are shown in histogram format. **c**, Normalized difference HE-LE for the absolute-valued trend’s first derivative magnitude and the difference in percentage for the threshold exceedance contextualized within the engaged home viewing population results. Observed values for all five matches are consistent with at-home VWs results.

### Engagement-related subthalamic activity is not explained by cardiac or movement signals

To exclude cardiac or gross motor artifacts (Methods), we performed an Ordinary Least Squares (OLS)-based variance decomposition of P-range amplitude in a subset of patients (*n* = 4; NWKb, NWKc, NWKd, NWKg) with concurrent biometric recordings (chest-mounted ROOTi-RX, Methods). Analysis of 5-6 days of continuous cardiac and kinematic data (Methods; fig. 8) confirmed that a substantial portion of the observed neural changes could not be explained by these factors.

P-range amplitude variance was partitioned into five components: “heart rate (HR)-unique”, “non-HR cardiac”, “kinematic-unique”, “shared” (cardiac + kinematic overlap), and “residual” (non-cardiac, non-kinematic). HR was used as the primary proxy for rate-dependent cardiac artifact contamination (Methods), with “HR-unique” representing the cardiac artifact upper bound. Non-HR autonomic signals (RMSSD, SDNN, LF/HF, EDR; Methods) are considered within the “non-HR cardiac” variance component, representing the biological lower bound. The “kinematic-unique” variance component is based on the signal magnitude area (SMA) feature quantifying movement-related P-range amplitude modulation (Methods). “HR-unique” variance accounted for 0.0-11.1% of total P-range amplitude variance across patients and hemispheres (median 2.1%). “Non-HR cardiac” explained an additional 1.3-9.4% of variance after HR removal, confirming genuine neural-cardiac coupling. “Kinematic-unique” variance was negligible in 7 of 8 patient-hemisphere combinations (range 0.0-3.4%, median 0.2%). The “residual” variance unexplained by cardiac or kinematic signals ranged from 68.5% to 97.2% (fig. 8a). Granger causality confirmed that SMA did not predict future P-range amplitude in 3 out of 4 patients. Conversely, P-range amplitude predicted future HR in 3 of 8 patient-hemisphere combinations (“neural→cardiac” dominant directionality). This indicates that part of the “HR-unique” variance reflects genuine neural-to-cardiac coupling rather than contamination (table Supplementary Information 4; Methods). Day-night modulation of the HR-unique component in one patient (NWKd Ch1: 6.2% daytime vs 17.8% nighttime) additionally indicated state-dependent biology within this estimate (table Supplementary Information 5).

A total of eight VWs with concurrent neural, cardiac, and kinematic recordings were captured within the 5-6 days of continuous monitoring (NWKb, *n* = 6 viewings; NWKc, *n* = 2 viewings; Methods), allowing assessment of cardiac and motor contributions to P-range amplitude modulation across engagement levels (HE vs LE). Within VWs, “kinematic-unique” contributions remained similarly small (0.0-3.9%). In-match HE-LE comparisons showed that gross trunk movement (SMA; Methods) did not consistently track engagement. The direction of SMA changes was inconsistent (Cohen’s d ranged from −0.45 to +0.68), with a pooled effect near zero (*d* = +0.023; fig. 8b). At the population level, HE-LE changes in P-range amplitude did not correlate with HE-LE changes in SMA (cross-viewing Spearman correlation; left: *ρ* < 0.01; right: *ρ* = 0.05; *n* = 8 viewings). This confirms that neural engagement modulation was decoupled from movement changes. Both P-range amplitude and time derivative effects were uncorrelated with HR effects (ΔP-range median vs ΔHR: left *ρ* = +0.10, right *ρ* = −0.13; Δ|*dP* −*range/dt*| vs ΔHR: left *ρ* = +0.25, right *ρ* = +0.41; all n.s.). Notably, cross-viewing correlation between P-range amplitude and HR Cohen’s d values was negative in the left hemisphere (*ρ* = −0.71, *p* = 0.047) (table Supplementary Information 5). Even after controlling for concurrent HR and movement (partial Spearman correlation, HR and SMA as covariates; Methods), P-range amplitude remained significantly correlated with engagement in 7 of 8 viewings (median partial *ρ* = +0.22; fig. 8c; table Supplementary Information 5). This indicates that P-range amplitude carried information beyond both cardiac and kinematic signals. Furthermore, adding P-range amplitude to a logistic regression model significantly improved engagement classification in 7 of 8 viewings, even when HR and movement were already included (likelihood-ratio test, *p* < 0.05; median ΔAUC= +0.048; maximum ΔAUC= +0.208 for NWKb Fiorentina-Como Ch2; fig. 8d; table Supplementary Information 5). Finally, engagement variance decomposition (Methods) revealed that the unique contribution of P-range amplitude to engagement prediction (median 3.1%) exceeded that of HR (median 0.9%) and of movement (median 1.9%). This confirms that P-range amplitude is not redundant but an independent source of engagement information. While the 95% CIs were broad due to the limited duration of HE/LE intervals, P-range amplitude consistently provided non-redundant information (table Supplementary Information 5). The single viewing in which none of the three analyses detected a significant P-range amplitude contribution in either hemisphere was Cremonese-Inter (patient NWKc). Notably, this viewing had been classified as neutral based on the post-viewing interview: while isolated moments captured the patient’s attention (defining brief HE intervals), the patient reported no sustained emotional engagement with the match (table Supplementary Information 2).

This triple dissociation (i.e., neural, cardiac, and kinematic signals each carrying independent engagement information), coupled with left-lateralized neural dynamics, indicates that P-range amplitude modulation reflects genuine neural processes not reducible to concurrent cardiac or kinematic changes (Discussion).

## Discussion

The basal ganglia are a functional brain node whose activity dynamically influences cortical areas involved in motor, associative, and limbic processing. Leveraging STN dynamics for aDBS requires real-time decoding of complex and context-dependent activity in real-world settings.

Our results show that the low-frequency (alpha-beta range) STN-LFP components dynamically track behavioral engagement during ecologically valid, prolonged experiences such as live sport match viewing (Fig. 1). Specifically, engagement was associated with a significant increase in P-range amplitude, not observed during neutral viewing at home (Fig. 2). Faster fluctuations were additionally coupled with salient in-match events, both at home (Fig. 3) and at the stadium (Fig. 4). These findings support the notion that STN activity encodes multidimensional information integrating emotional, cognitive, and contextual factors. Accordingly, they support an expanded functional role beyond motor processing and point to context-aware and personalized aDBS strategies.

Our study leveraged chronic bilateral STN-LFP monitoring using the AlphaDBS system[16, 17, 34], enabling continuous, real-world recording of STN activity over extended periods (Fig. 1a; fig. Extended Data 1; tables Supplementary Information 1, Extended Data 1). The dual-resolution strategy, combining high-temporal-resolution P-range amplitude sampling (1-min; Fig. 1c) with lower-resolution full-spectrum recordings (10-min; Fig. 1d; fig. Extended Data 4), allowed us to capture both rapid fluctuations and long-term trends in neural dynamics. Importantly, this dataset represents one of the most comprehensive collections of chronic STN recordings during sustained naturalistic behavior in PD patients[35], providing unprecedented insights into neural dynamics across multiple temporal scales in authentic experiential contexts.

In contrast to laboratory paradigms, which rely on brief, isolated emotional stimuli lasting seconds, our approach captures sustained engagement unfolding over minutes to hours, characterized by anticipation, uncertainty, and evolving emotional states. Standard paradigms such as the International Affective Picture System (IAPS) images[36] or Montreal Affective Voices[37] fail to model the temporal continuity and cognitive-emotional integration typical of long lasting real-world experiences[38]. Moreover, laboratory conditions minimize ecological validity by isolating participants from motor, social, and contextual influences[39], whereas our paradigm incorporates these factors naturally.

Several principles guided our ecological approach to biomarker discovery. First, paradigms should induce sustained cognitive-emotional engagement over clinically relevant timescales (minutes to hours). Second, they should include varying levels of motor activity to disentangle motor and non-motor contributions to STN signals. Third, engagement should vary both between individuals (enabling population-level comparisons between engaged versus neutral states) and within individuals over time (enabling identification of temporal signatures accompanying engagement fluctuations). Fourth, salient events embedded within prolonged experiences should allow investigation of multi-scale encoding. Finally, paradigms must be meaningful to participants, ensuring genuine emotional investment rather than artificial task compliance[40].

Engaged VWs at home showed a sustained bilateral increase in P-range amplitude relative to matched baseline BD and control CP (Fig. 2; table Supplementary Information 3), consistent with a tonic shift in brain state. The specificity to engaged viewing was supported by post-viewing interviews (fig. Extended Data 3; table Extended Data 3 for the interview protocol; table Supplementary Information 2 for the transcriptions), further suggesting that our finding reflects a neural correlate of sustained cognitive-emotional engagement. This effect is unlikely to be driven by confounds such as circadian variation, sleep-wake cycles17 or slow pathological drifts[41] (fig. Extended Data 2; Methods). Importantly, the parallel increase in stimulation current during engaged viewing (< 0.1 mA; fig. Extended Data 7) confirms the clinical relevance of these engagement-related neural dynamics, as the aDBS system interpreted elevated P-range amplitude as requiring increased stimulation.

Using an additive decomposition approach (Methods), we demonstrated that engagement modulates STN activity across multiple temporal scales (Fig. 1e). While P-range amplitude changes were bilateral (Fig. 2), time derivatives were predominantly left-lateralized (Fig. 3c-e), indicating a functional dissociation between sustained state encoding and dynamic temporal processing across hemispheres. At slower timescales, time derivatives suggested gradual transitions between LE and HE states (Fig. 3b; fig. Extended Data 5), reflecting sustained and non-linear reconfiguration of STN activity. These dynamics likely capture overlapping neural processes underlying naturalistic engagement, consistent with the structure of real-world paradigms in cognitive neuroscience[42]. In this context, sports matches unfold over extended periods with a hierarchical narrative structure[43], where anticipation builds progressively as outcome uncertainty (e.g., tight scores) converges with temporal proximity to resolution (e.g., decisive points), generating sustained anticipatory states lasting minutes. Importantly, stimuli were identical across patients, ensuring reliability and enabling inter-patient comparisons despite the naturalistic setting. At faster timescales, salient in-match events were temporally coupled with P-range amplitude local extrema (Fig. 3d), yet without significant maximum variation-at-events (Methods), suggesting sustained engagement rather than reactive responsivity to individual events. The presence of both local maxima and minima suggests that engagement is not solely encoded by P-range amplitude changes but reflects phase-dependent oscillatory dynamics, potentially involving phase resetting or synchronization mechanisms within cortico-basal ganglia networks[44, 45]. Analysis of inter-versus intra-individual variability (Methods) further supports this interpretation. While P-range amplitude exhibited high inter-individual variability (ICC*>* 86%), time derivatives showed markedly lower variability across participants (ICC< 3%), indicating greater generalizability. This property makes time derivative-based features attractive candidates for adaptive algorithms, potentially reducing the need for extensive patient-specific calibration.

The stadium recordings (Fig. 4) provided a naturalistic framework to disentangle engagement from motor activity. Despite increased movement (fig. Extended Data 6), which can suppress STN P-range amplitude[14, 46], time derivative dynamics and event-related coupling were preserved (Fig. 4c), whereas sustained P-range amplitude increases were not (Fig. 4b). This dissociation suggests that temporal features encode engagement independently of motor state, consistent with a multiplexed coding scheme in which P-range amplitude, time derivatives, and local extrema convey distinct yet complementary information channels. These observations align with evidence implicating the STN in cognitive and affective processes beyond motor control[47, 48]. Importantly, our findings extend this framework to naturalistic conditions, demonstrating that engagement-related information remains encoded even when motor influences dominate P-range amplitude.

Current aDBS systems rely primarily on P-range amplitude thresholds optimized for motor symptoms control[49]. In our dataset, this approach produced only modest stimulation amplitude changes during engagement (< 0.1 mA). Although it is not possible to determine how neural activity would have evolved in the absence of this adjustment, the observed variation, while statistically significant and potentially biologically relevant, remained below the threshold of clinical (motor) relevance. Context-aware algorithms incorporating multi-scale temporal features may better distinguish pathological activity from adaptive neural dynamics, thereby improving both efficacy and specificity.

Several limitations should be considered. The cohort comprised only eight patients with varying degrees of emotional involvement (e.g., being highly engaged supporters or less emotionally invested). For simplicity, we adopted a dichotomous classification of engaged versus neutral states based on retrospective interviews, which may mask interindividual variability. Additionally, in 8 of 62 monitored periods, only unilateral data (Ch1/Left) were available, as bilateral research recording capabilities had not yet been enabled at the time of the monitoring. Recording interruptions occurred in some sessions, primarily during device charging cycles; however, these interruptions did not affect therapeutic stimulation delivery, which continued uninterrupted. Despite these occasional interruptions, which represent an expected limitation of chronic neural recordings in real-world conditions[50], the dataset provided sufficient data to support robust conclusions. Multimodal recordings in a subset of participants (Methods) indicated that most variance in the P-range amplitude was not explained by cardiac or kinematic factors (fig. 8; tables Supplementary Information 4, Supplementary Information 5), supporting a predominantly neural origin; however, residual confounds cannot be fully excluded (Methods). Future longitudinal home-monitoring protocols integrating clinical diaries with multiple wearable measurements synchronized with STN recordings will enable supervised learning approaches for state classification.

Together, these findings redefine the functional role of the STN within neuromodulation, shifting the perspective from a symptom-centric controller of pathological oscillations to a dynamic, multimodal integrator of behaviorally relevant brain states. By demonstrating that engagement-related information is encoded across multiple temporal scales and remains detectable even under varying motor conditions, this work establishes a principled framework for next-generation aDBS systems that operate in the full complexity of real-world environments. In this view, effective neuromodulation will not simply suppress aberrant activity but will continuously interpret and adapt to the patient’s evolving cognitive, emotional, and motor state space, ultimately enabling truly personalized, context-aware brain-machine interfaces that align therapeutic intervention with the lived experience of the individual.

## Methods

### Study Design and Data Collection

Eight patients with idiopathic PD (Hoehn and Yahr stage 2 of 5 on optimal medical treatment) agreed to participate in this study. They constitute part of the same cohort of patients reported in our previous studies, as detailed in table Extended Data 2. Patients underwent bilateral STN DBS implantation using either Medtronic 3389 leads or directional Abbott 6172 leads (fig. Extended Data 1; table Supplementary Information 1) for clinical indications and received the AlphaDBS (Newronika SpA) IPG as part of a clinical trial (Trial registration number NCT04681534[51]). The recordings in this study were performed after completion of the clinical trial in patients who preferred aDBS at the end of the clinical study. This study was approved by the local Ethics Committee (approvals 165-2020 and 93-2023bis), conformed to the Declaration of Helsinki, and all patients provided written informed consent. Demographic and clinical characteristics, including Levodopa Equivalent Daily Dose (LEDD), are provided in table Extended Data 1. Importantly, throughout each monitoring period of this study, patients’ pharmacological therapy remained unchanged, with medication intake maintained at the same scheduled times. Patients were clinically stable during each monitoring period, and clinical assessment was performed using the Unified Parkinson’s Disease Rating Scale (UPDRS) Part III (motor section) within one month of monitoring.

Patients watched live, high-stakes tennis matches (Roland Garros 2025 Final, Wimbledon 2025 Final, US Open 2025 Final, multiple ATP Finals 2025 matches, and Australian Open 2026 Semifinals and Final) and/or soccer matches (UEFA Champions League 2025 Semifinal 2nd leg and Final, Serie A 2025/2026 and Coppa Italia 2025/2026 matches) at home, quietly seated in front of the television to minimize movement-related confounds during cognitive-emotional engagement with the match (Fig. 1a,b). A total of 57 home viewings were collected. One patient (NWKc) additionally watched five matches at the stadium (UEFA Champions League 2025 Semifinal 2nd leg and Final, Serie A 2025/2026 1st round Derby della Madonnina, Serie A 2025/2026 1st round FC Internazionale Milano vs Bologna FC, and Serie A 2025/2026 2nd round Derby d’Italia), where the context varied substantially from the home environment due to the different kind of stimuli and the different level of motor activity. These viewings were analyzed separately as a test case.

Bilateral STN-LFPs amplitude was chronically recorded (left hemisphere: channel Ch1; right hemisphere: channel Ch2; Fig. 1c) using the AlphaDBS device[52, 53]. Recordings captured a personalized frequency range within the alpha-beta band (P-range, within 5-23 Hz) at 1-min temporal resolution. For each patient, the P-range is centered on the most prominent peak in the alpha-beta band correlating with motor symptoms and serving as the patient-specific clinical biomarker. Full amplitude spectra (5-34 Hz) were simultaneously recorded at 10-min resolution (Fig. 1d). For 8 of 62 monitored periods, only unilateral (Ch1/Left) data were available, as bilateral research recording had not yet been enabled on those dates (table Supplementary Information 3).

### Monitoring periods and analysis windows

Recordings acquired during each match were compared with those acquired within a monitoring period spanning ±3-4 consecutive days (7-10 days total) (fig. Extended Data 2). This duration was selected to minimize potential contamination from pathological long-period oscillations in the P-range while providing adequate baseline data. To analyze the ATP Finals viewings of patient NWK, which occurred on multiple days within a single week, the preceding week served as control period. For Australian Open and Serie A viewings (patients NWKb and NWKc), which occurred within two to three days, controls were adequately treated to ensure that no other viewing period was included (i.e., minutes within viewing periods not of interest were not retained as controls).

Within each monitoring period, analyses focused on time series (of both 1-min and 10-min resolution, see below the Signal Processing section) during viewing window (VW), the baseline day (BD), and the control period (CP). VW time series consist of time points within 5 min pre-match to 5 min post-match. BD time series consist of time points from the match day, external to the VW. CP time series consist of time points from the rest of the days within the monitoring period synchronous to the VW. In both BD and CP, nighttime [10:00pm - 6am] was excluded to avoid bias in our results due to a lower STN P-range amplitude during night compared to daytime. In VWs, periods in which patients explicitly reported to be asleep were removed.

In addition, we identified 12 non-viewing windows temporally aligned with major matches (two soccer matches: UEFA Champions League 2025 Semifinal 2nd leg and Final; two tennis matches: Roland Garros 2025 Final, Wimbledon 2025 Final), during which patients reported engaging in activities other than match viewing.

### Engagement evaluation

At enrollment, five patients were identified as supporters: three of FC Internazionale Milano (NWKa, NWKc, NWKg), one of Juventus FC (NWKd), and one of Como 1907 and tennis player Jannik Sinner (NWKb). The remaining patients (NWKe, NWKf, NWKh) expressed general interest in sports but were not supporters of the specific teams or players competing in the monitored matches.

Post-viewing semi-structured interviews conducted 2-7 days after each match classified VWs as engaged (*n* = 29) or neutral (*n* = 17) based on subjective interest. The interview protocol consisted of an initial free-recall narrative (monologue) of viewing experience followed by five structured questions examining: (1) viewing context and continuity, (2) temporal dynamics of emotional states across match segments, (3) specific discrete emotions (anger, sadness, confidence, attention), (4) expectation formation and outcome acceptance, and (5) comparative engagement relative to previous viewing experiences. Interviews were conducted in participants’ native language (Italian) and responses were transcribed (tables Extended Data 3, Supplementary Information 2).

Engaged VWs elicited spontaneous, detailed match narratives with pronounced affective responses, particularly from team supporters who provided extended, unprompted recollections. Non-supporters involved in engaged VWs demonstrated comparable event recall but primarily through directed responses rather than voluntary commentary. Neutral VWs showed minimal specific event recall despite confirmed viewing, indicating passive attention. This distinction was quantifiable through recall granularity: viewers involved in engaged VWs reproduced detailed, temporally-structured accounts of critical match moments and associated emotional states, whereas viewers involved in neutral VWs could not retrieve specific details when prompted (table Supplementary Information 2).

### In-Match Engagement and Event Annotation

For each engaged VWs, we further partitioned the recording into higher engagement (HE) and lower engagement (LE) intervals based on temporal details in interview responses. HE intervals encompassed match periods explicitly and spontaneously recalled as eliciting heightened interest, excitement, or emotional investment. These intervals were defined by the onset or offset of specific in-game events (see below), as goals in soccer or break-points in tennis. LE intervals comprised remaining match periods not specifically highlighted during interview recall. This temporal partitioning enabled within-session comparison of neural signatures corresponding to subjectively distinct engagement states.

Times of key events within each match were identified at 1-min resolution using online sports newspapers (e.g., “Corriere dello Sport”, “OA sport”, “Gazzetta dello Sport”, and “Euro Sport”) and validated through visual inspection of match recordings. For tennis matches, reported times were used without adjustment. For soccer matches, reported times were used directly for stadium viewings, while 1 min was added for home viewings to account for television broadcast delays[54].

For home viewings, analysis was restricted to score-related events (goals, penalties, break points, set points, match points), as these represent unambiguously salient in-match moments. Non-score events were excluded because, without direct observation of the patient, there was no means to determine which of those events the patient was actively following or emotionally responding to. For stadium viewings, where the richer in-person context provided greater certainty about patient involvement, the event set was extended to include near-misses, disciplinary actions (yellow cards), and substitutions. For the ATP Finals 2025 Final, NWKb provided during the post-match interview a personal annotation of subjectively salient moments; this was included as an additional event set exclusively for that viewing, as it uniquely allowed extending the analysis beyond score-related events for a home VW.

### Signal processing

STN-LFP data were extracted from two time series acquired separately52: (i) the 1-min resolution amplitude in the personalized P-range provided by the AlphaDBS device and driving stimulation delivery (Fig. 1c; fig. Extended Data 2), and (ii) the 10-min resolution amplitude across standard frequency bands (theta: 5-8 Hz; alpha: 8-12 Hz; low-beta: 12-20 Hz; high-beta: 20-30 Hz; early-gamma: 30-34 Hz) derived from the amplitude spectra provided by the device (Fig. 1d; fig. Extended Data 4a,b). At each time point, amplitude was computed as the sum of STN-LFP amplitude across frequency bins within the specified band, normalized by the sum of amplitude across 5-34 Hz to account for inter-individual differences in signal magnitude. In addition, the recorded bilateral stimulation current, delivered to the STN through a linear proportional algorithm17,52, was directly provided by the device with 1-min resolution. Median amplitudes for each VWs and corresponding BD and CP were computed and compared.

For 1-min resolution P-range amplitude during engaged VWs, we additionally performed an in-match temporal analysis (Fig. 1e). Two engaged VWs (patient NWKc, US Open 2025 Final and ATP Finals 2025 Sinner-Zverev) were excluded from this analysis due to recording gaps of more than ten consecutive minutes caused by device charging. We adopted an additive decomposition model looking at the engaged VW signal *Y* [*t*] as the sum of trend *T*[*t*] and detrended *D*[*t*] components. Trend component was computed at each minute t of engaged VWs as the centered average of a 30-min long period across the initial raw time series, while the detrended component resulted from the difference of the original time series and its trend. To mitigate border effects, to extract the trend component we expanded each engaged VW by considering points within 30 min pre-match to 30 min post-match.

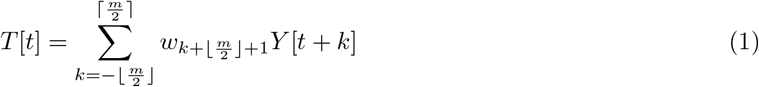

where *m* = *p* + 1 is the length of the sliding window filter when the period *p* is odd, 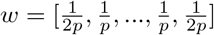 is the coefficient array of length *m*.

Trend’s first and second time derivatives were computed through the Savitzky-Golay Filter, consisting of local polynomial fitting to the signal within a sliding window where polynomial coefficients were found via least squares optimization (Fig. 3b). We used a 15-min long sliding window, appropriate for adequate smoothing of our 1-min resolution signal allowing to test polynomial up to the 13^*th*^ order.

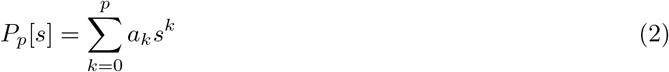

where 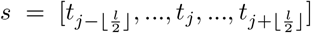 array of time points, *l* is the length of the sliding window and *a*_0_, *a*_1_, …, *a*_*p*_ are the coefficients:

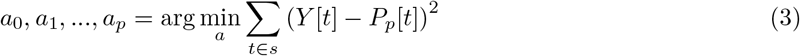

The optimal order *p*\* of the fitting polynomial for each VW was computed by testing odd values from 3 to 13 and selecting the one that maximized the composite score defined as the sum of the fitting quality score (based on *R*^2^), the error score (based on MSE, inverted), the first time derivative discrimination score and the second time derivative discrimination score (both based on the median magnitude of trends’ time derivatives).

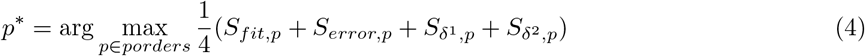

where

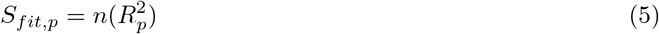

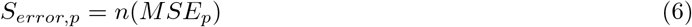

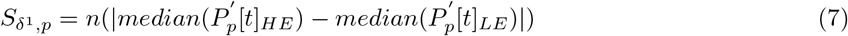

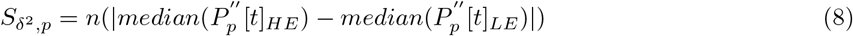

with

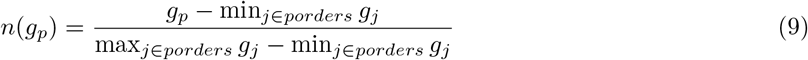

Optimal polynomial orders ranged from 9 to 13 across viewings.

First time derivative quantified the instantaneous rate of change in the trend signal, reflecting the velocity of neural state changes. Second time derivative quantified the acceleration or curvature of trend dynamics. Aiming at characterizing HE and LE windows, we compared the median magnitudes of the absolute-valued time derivatives and the percentage of time points where each time derivative exceeded its own 75^*th*^ percentile computed across the entire VW.

For the detrended component (Fig. 3d), local maxima and minima (extrema) were identified with two threshold values (0.0 and 0.5) to assess sensitivity to small fluctuations. We quantified two complementary metrics of event-related modulation: (i) extrema-near-events proportion (fraction of events with a P-range amplitude extremum within ±1 min), and (ii) maximum variation-at-events (largest P-range amplitude variation between event times and adjacent time points within ±1 min of the event).

Stimulation current time series were extracted during VW, CP, BD windows and analyzed using identical statistical frameworks. Analysis focused on median current values per hemisphere per viewing session to parallel the P-range amplitude analysis structure.

### Cardiac and kinematic artifact control analysis

For a subset of patients (*n* = 4; NWKb, NWKc, NWKd, NWKg) we were able to perform 5-6 days of concurrent cardiac and kinematic monitoring (fig. 8, table Supplementary Information 4). This enabled direct quantification of the cardiac and kinematic contributions to P-range amplitude variance. Cardiac activity features (heart rate HR, root mean square of the successive differences RMSSD, standard deviation of normal-to-normal intervals SDNN, low frequency - high frequency LF/HF ratio, ECG derived respiration EDR) and tri-axial chest accelerometry (31.25 Hz sampling rate) were recorded by a chest-mounted wearable device (ROOTi-RX, Rooti Labs), resampled at 1-min resolution and synchronized with the neural recordings by the internal clocks of the two recording devices.

Signal magnitude area (SMA) was computed from raw data as the sum of absolute sample-to-sample differences in acceleration (i.e., as the incremental activity count IAC[55, 56]) across the three axes within each minute:

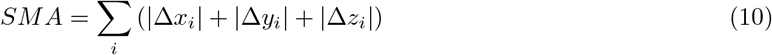

where Δ*x*_*i*_ = *x*(*t*_*i*_) − *x*(*t*_*i*−1_) represents the change in acceleration along axis *x* between consecutive samples.

Unlike the conventional SMA (i.e., the integral of absolute acceleration), this formulation removes the static gravitational component through differencing, isolating dynamic acceleration from trunk movement and postural changes without requiring gravity calibration55,56. Vector magnitude (VM), averaged per minute, was retained as a complementary measure of overall acceleration intensity.

HR was treated as the primary proxy for cardiac artifact contamination; as autonomic indices derived from heart rate variability (RMSSD, SDNN, LF/HF, EDR) are more likely to reflect regulatory processes governing the heart rather than the electrical contamination itself, they were treated exclusively as biologically informative signals (Discussion). A limitation of this approach is that it addresses rate-dependent contamination only; changes in QRS morphology accompanying sympathetic activation could introduce additional artifact variance not captured by heart rate.

We decomposed P-range amplitude variance into five components using OLS regression models on the full monitoring period (fig. 8a). Each OLS model provided a coefficient of determination R^2^ quantifying the total variance in P-range amplitude explained by that predictor set; unique variance components were then computed as differences between nested models, isolating the variance exclusively attributable to each predictor group beyond what the remaining predictors already explain. The full model regressed P-range amplitude on all cardiac and kinematic predictors simultaneously:

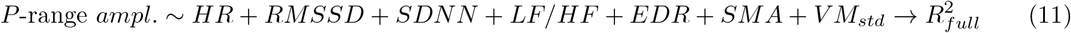

Reduced models omitted either HR alone, all kinematic predictors, or all non-HR cardiac predictors:

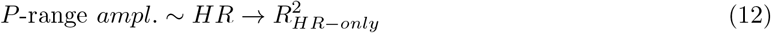

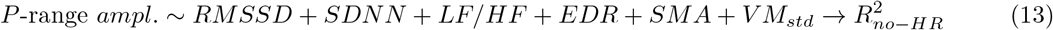

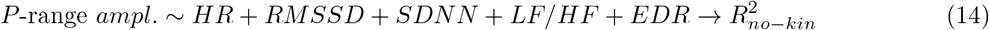

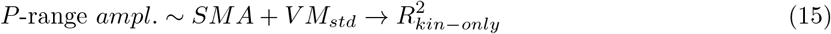

Each variance component was then computed as the incremental R^2^ lost when removing the corresponding predictor group from the full model:

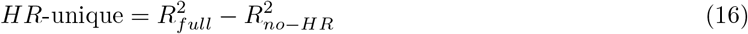

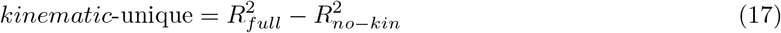

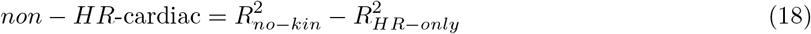

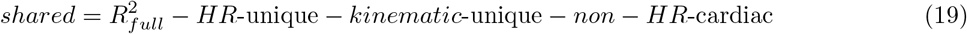

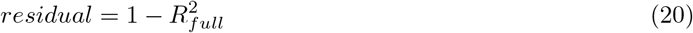

The five components sum to unity. “HR-unique” variance provides an upper bound on rate-dependent artifact, since it also captures genuine HR-mediated biological coupling. “Kinematic-unique” quantifies movement-related P-range amplitude modulation. “Non-HR cardiac” variance provides a lower bound on genuine neural-cardiac coupling. “Residual” represents neural variance unrelated to any peripheral signal. 95% CI were estimated via bootstrap (2000 resamples preserving within-minute pairings). Granger causality analysis tested predictive directionality between HR and P-range amplitude, and between SMA and P-range amplitude, after confirming stationarity (Augmented Dickey-Fuller ADF and Kwiatkowski-Philips-Schmidt-Shin KPSS tests with detrending for the cardiac signal; table Supplementary Information 4). The same decomposition was applied separately to daytime (6am-10pm) and nighttime (10pm-6am) periods (table Supplementary Information 4).

To test for potential cardiac or kinematic contamination of P-range amplitude, we compared P-range amplitude, HR, and SMA across HE and LE intervals within each viewing session (*n* = 8 viewings, six from NWKb and two from NWKc; Mann-Whitney U tests). Effect sizes were quantified using Cohen’s d and rank-biserial correlation (fig. 8b; table Supplementary Information 5). Cross-viewing Spearman correlations tested whether HE-LE effects in P-range amplitude and in trend time derivative magnitude covaried with HE-LE effects in HR and SMA across viewings. A separate cross-viewing Spearman correlation was computed between Cohen’s d values of P-range amplitude and HR HE-LE effects (table Supplementary Information 5).

Three complementary in-match analyses then quantified the extent to which engagement is encoded in P-range amplitude after controlling for cardiac and kinematic confounds by removing linear rank influence of HR and SMA. First, partial Spearman correlations measure the association between minute-by-minute P-range amplitude and the binary HE/LE engagement label (fig. 8c). Second, nested logistic regression models were compared: a confound model (*M*_0_ : *engagement* ~ *HR*+*SMA*) and a full model (*M*_1_ : *engagement* ~ *HR* + *SMA* + *P* − *range*), with all predictors standardized; the likelihood-ratio test (*χ*^2^, dof = 1) assessed whether P-range significantly improved model fit, and incremental 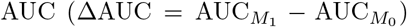 quantified the discriminative improvement conferred by its inclusion (fig. 8d). Third, to quantify the relative magnitude of each predictor’s contribution, an engagement variance decomposition partitioned HE/LE label variance into “P-range-unique”, “HR-unique”, “SMA-unique”, “shared”, and “residual” components via incremental OLS *R*^2^, with 95% bootstrap CI (2,000 iterations).

### Statistical Analysis Framework

#### Statistical tests

Per-match, per-hemisphere VW-CP and VW-BD P-range amplitude comparisons used the Mann-Whitney U test with Benjamini-Hochberg false discovery rate (FDR) correction for multiple comparisons (Fig. 2a; table S6); the same test was applied to HE-LE comparisons of P-range amplitude, HR, and SMA within VWs (fig. 8b; table Supplementary Information 5). Population-level analyses employed the Wilcoxon signed-rank test on paired medians:

- (VW vs. CP; VW vs. BD) for engaged, neutral, and non-viewing categories, both for P-range amplitude (Fig. 2b, 2c; fig. Extended Data 3) and stimulation current (fig. Extended Data 7);
- (HE vs. LE) for absolute-valued trend time derivatives and proportion of time points exceeding the 75^*th*^ percentile (Fig. 3c, fig. Extended Data 5b).

Effect sizes were quantified using Cohen’s d, with thresholds of |*d*| ≥ 0.7 for large effects, |*d*| ≥ 0.3 for medium effects, |*d*| ≥ 0.1 for small effects, |*d*| < 0.1 for negligible effects (Fig. 2b, 2c, 3c, 4b, 4c; fig. Extended Data 3).

Because engagement could only be assessed during viewing, a full 2*x*2 factorial design (two-way ANOVA) was not possible. Consequently, each category was analyzed independently rather than testing for interaction.

#### Linear Mixed Effects modeling

To account for repeated engaged VWs within patients (*n* = 6 patients, 1-6 VWs per each patient), we employed linear mixed-effects models using the ‘statsmodels’ package in Python. Models included condition (target, control) as a fixed effect and patient as a random intercept, fitted via restricted maximum likelihood (REML). This approach addresses the non-independence of repeated measurements within patients while leveraging all available data from multiple VWs. This framework was applied to: (i) P-range amplitude (Fig. 2b); and (ii) absolute-valued trend time derivatives and proportion of time points exceeding the 75^*th*^ percentile (Fig. 3c; fig. Extended Data 5b). The general model structure was:

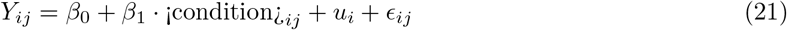

where *Y*_*ij*_ is the outcome measure (i.e., P-range amplitude or trend derivative magnitude) for patient *i* in episode *j, β*_0_ is the population intercept, *β*_1_ is the fixed effect of *< condition >*, 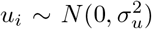 is the patient-specific random intercept, 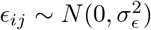 is the residual error term.

We first validated whether CP and BD were statistically equivalent (*< condition >*_*ij*_= *CP, BD*_*ij*_), by fitting a preliminary model comparing only these two reference conditions. Upon reaching *p* > 0.05 in this comparison (i.e., statistical equivalence), we pooled both CP and BD into a single pooled reference category for the primary analysis, maximizing statistical power.

We then tested the fixed effect of condition on our neural data:

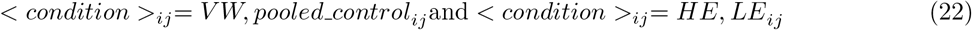

where the fixed effect coefficient *β*_1_ represents the population-level difference in the outcome measure *Y*_*ij*_, accounting for patient-specific baselines through the random intercept *u*_*i*_. All models were fitted using the Powell optimization algorithm and verified for convergence, normality of random effects, and homoscedasticity of residuals.

For each model, we calculated the intraclass correlation coefficient ICC to quantify between-patient variance and Cohen’s *d* for effect sizes:

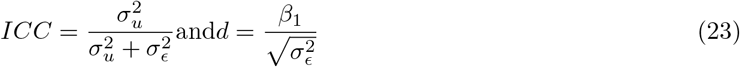

where 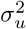 is the between-patient variance, 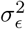 is the within-patient residual variance. High ICC values (*>* 50%) indicate substantial individual differences in baseline levels, justifying the mixed-effects approach. Statistical significance was assessed using two-tailed z-tests (*α* = 0.05), with Benjamini-Hochberg FDR correction applied for multiple comparisons (i.e., multiple frequency bands).

#### Event-related signal analysis

Statistical significance of extrema-near-events and maximum variation at-events in the detrended signal was assessed via non-parametric permutation testing with 10,000 iterations. For each iteration, event times were randomly shuffled, and both metrics were recalculated to generate null distributions (Fig. 3e). One-tailed (right-tailed) tests were applied to assess whether observed peri-event amplitude associations (i.e., both extrema-near-events and variation-at-events) exceeded chance levels. The same permutation testing framework was applied to the five stadium viewings from NWKc for the extremanear-events metric (*n* = 5 VWs) and to the three stadium viewings with available right-STN data for the maximum variation-at-events metric (*n* = 3 VWs), reported descriptively given the limited sample (Fig. 4).

This approach increased statistical power to detect effects in the predicted direction (i.e., events temporally aligned with neural extrema) compared to two-tailed tests. Statistical power for permutation tests was estimated post-hoc using parametric approximations based on the observed effect size, sample size, and significance level (*α* = 0.05, one-tailed). For each analysis, we calculated retrospective power as

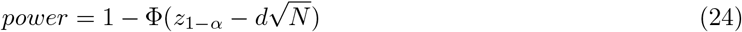

where *z*_1−*α*_ is the critical value (1.645 for *α* = 0.05, one-tailed), Φ(·) is the standard normal CDF, *d* is the observed Cohen’s *d* effect size, and *N* is the number of events. Minimum detectable effect sizes (*d*_*MDES*_) and required sample sizes for 80% power were calculated as

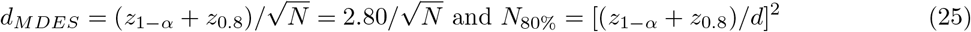

Due to the limited number of events, individual matches provided insufficient power (estimated 15-45%) to detect medium effects (*d* ≈ 0.5). To address this limitation, we aggregated z-scored normalized data across multiple VWs within individual patients, under the hypothesis of similar within-patient engagement for all engaged VWs recorded (Figure 3d, 4d). With this constraint, this kind of analysis resulted feasible (power *>* 80% for detecting the observed effect size) only for three patients: (i) NWKb with *n* = 12 engaged VWs, NWKc with *n* = 8 engaged VWs (5 of them at the stadium), and NWKd with *n* = 5 engaged VWs (see table Supplementary Information 3 to retrieve the corresponding matches).

## Data Availability

The datasets presented in this article are not readily available because LFP recorded with the AlphaDBS device cannot be deposited in a public repository as they can be traceable to the identity of the subject. Requests to access the datasets should be directed to IUI.

## Acknowledgments

We are grateful to many colleagues for their help in this line of research. In particular, we would like to thank: Alessandra Ranghetti, Salvatore Bonvegna of the Parkinson Institute of Milan, ASST G.Pini-CTO; Nicolò Pozzi and Ibrahem Hanafi of the University Hospital of Wuerzburg; Jacopo Carpaneto and Michele Valla of the Biorobotics Institute of the Sant’Anna School of Advanced Studies.

## Author contribution

SF: Conceptualization, Formal Analysis, Investigation, Methodology, Software, Writing-original draft. LC: Data curation, Investigation, Methodology, Writing-review and editing. FL: Data curation, Writing-review and editing. CP: Data curation, Investigation, Methodology, Writing-review and editing, Funding acquisition, Resources, Supervision. IUI: Conceptualization, Formal Analysis, Investigation, Methodol ogy, Writing-original draft, Funding acquisition, Resources, Supervision. AM: Conceptualization, Formal Analysis, Investigation, Methodology, Writing-original draft, Funding acquisition, Resources, Supervision.

## References

[1] Zhu, J. et al. Temporal trends in the prevalence of parkinson’s disease from 1980 to 2023: a systematic review and meta-analysis. The lancet Healthy longevity 5, e464–e479 (2024).

[2] de Bie, R. M. et al. Update on treatments for parkinson’s disease motor fluctuations–an international parkinson and movement disorder society evidence-based medicine review. Movement Disorders 40, 776–794 (2025).

[3] Oikonomou, P., Akhoundi, F. H., Olfati, N. & Litvan, I. Characteristics and mechanisms of cognitive impairment in parkinson disease. Nature Reviews Neurology 1–20 (2025).

[4] Den Brok, M. G. et al. Apathy in parkinson’s disease: a systematic review and meta-analysis. Movement Disorders 30, 759–769 (2015).

[5] Reijnders, J. S., Ehrt, U., Weber, W. E., Aarsland, D. & Leentjens, A. F. A systematic review of prevalence studies of depression in parkinson’s disease. Movement disorders 23, 183–189 (2008).

[6] Chaudhuri, K. R. & Schapira, A. H. Non-motor symptoms of parkinson’s disease: dopaminergic pathophysiology and treatment. The Lancet Neurology 8, 464–474 (2009).

[7] Drui, G. et al. Loss of dopaminergic nigrostriatal neurons accounts for the motivational and affective deficits in parkinson’s disease. Molecular psychiatry 19, 358–367 (2014).

[8] Pagonabarraga, J., Kulisevsky, J., Strafella, A. P. & Krack, P. Apathy in parkinson’s disease: clinical features, neural substrates, diagnosis, and treatment. The Lancet Neurology 14, 518–531 (2015).

[9] Deuschl, G. et al. A randomized trial of deep-brain stimulation for parkinson’s disease. New England Journal of Medicine 355, 896–908 (2006).

[10] Okun, M. S. Deep-brain stimulation—entering the era of human neural-network modulation. New England Journal of Medicine 371, 1369–1373 (2014).

[11] Krauss, J. K. et al. Technology of deep brain stimulation: current status and future directions. Nature Reviews Neurology 17, 75–87 (2021).

[12] Little, S. et al. Adaptive deep brain stimulation in advanced parkinson disease. Annals of neurology 74, 449–457 (2013).

[13] Little, S. & Brown, P. The functional role of beta oscillations in parkinson’s disease. Parkinsonism & related disorders 20, S44–S48 (2014).

[14] Cassidy, M. et al. Movement-related changes in synchronization in the human basal ganglia. Brain 125, 1235–1246 (2002).

[15] Tinkhauser, G. et al. Beta burst dynamics in parkinson’s disease off and on dopaminergic medication. Brain 140, 2968–2981 (2017).

[16] Arlotti, M. et al. Eight-hours adaptive deep brain stimulation in patients with parkinson disease. Neurology 90, e971–e976 (2018).

[17] Caffi, L. et al. Adaptive vs. conventional deep brain stimulation: one-year subthalamic recordings and clinical monitoring in a patient with parkinson’s disease. Bioengineering 11, 990 (2024).

[18] Prasad, A. A. & Wallén-Mackenzie, Å. Architecture of the subthalamic nucleus. Communications Biology 7, 78 (2024).

[19] Hammond, C., Bergman, H. & Brown, P. Pathological synchronization in parkinson’s disease: networks, models and treatments. Trends in neurosciences 30, 357–364 (2007).

[20] Serranová, T. et al. Topography of emotional valence and arousal within the motor part of the subthalamic nucleus in parkinson’s disease. Scientific Reports 9, 19924 (2019).

[21] Benis, D. et al. Subthalamic nucleus activity dissociates proactive and reactive inhibition in patients with parkinson’s disease. Neuroimage 91, 273–281 (2014).

[22] Jin, Y. et al. Altered emotional prosody processing in patients with parkinson’s disease after subthalamic nucleus stimulation. Neuropsychiatric Disease and Treatment 2965–2975 (2017).

[23] Pell, M. D. & Leonard, C. L. Processing emotional tone from speech in parkinson’s disease: a role for the basal ganglia. Cognitive, Affective, & Behavioral Neuroscience 3, 275–288 (2003).

[24] Ceravolo, L., Frühholz, S., Pierce, J., Grandjean, D. & Péron, J. Basal ganglia and cerebellum contributions to vocal emotion processing as revealed by high-resolution fmri. Scientific reports 11, 10645 (2021).

[25] Baumgartner, A. J., Hirt, L., Amara, A. W., Kern, D. S. & Thompson, J. A. Diurnal fluctuations of subthalamic nucleus local field potentials follow naturalistic sleep-wake behavior in parkinson’s disease. Sleep 48, zsaf005 (2025).

[26] van Rheede, J. J. et al. Diurnal modulation of subthalamic beta oscillatory power in parkinson’s disease patients during deep brain stimulation. npj Parkinson’s Disease 8, 88 (2022).

[27] Cagle, J. N. et al. Chronic intracranial recordings in the globus pallidus reveal circadian rhythms in parkinson’s disease. Nature Communications 15, 4602 (2024).

[28] Provenza, N. R. et al. Long-term ecological assessment of intracranial electrophysiology synchronized to behavioral markers in obsessive-compulsive disorder. Nature medicine 27, 2154–2164 (2021).

[29] Arnulfo, G. et al. Phase matters: a role for the subthalamic network during gait. PloS one 13, e0198691 (2018).

[30] Thompson, J. A. et al. Sleep patterns in parkinson’s disease: direct recordings from the subthalamic nucleus. Journal of Neurology, Neurosurgery & Psychiatry 89, 95–104 (2018).

[31] Lozano, A. M. et al. Deep brain stimulation: current challenges and future directions. Nature Reviews Neurology 15, 148–160 (2019).

[32] Ehgoetz Martens, K. A., Ellard, C. G. & Almeida, Q. J. Does anxiety cause freezing of gait in parkinson’s disease? Plos one 9, e106561 (2014).

[33] Falciglia, S. et al. Task-related biomarkers and technical developments for adaptive deep brain stimulation in parkinson’s disease. Brain Stimulation: Basic, Translational, and Clinical Research in Neuromodulation 18, 273 (2025).

[34] Isaias, I. U. et al. Case report: Improvement of gait with adaptive deep brain stimulation in a patient with parkinson’s disease. Frontiers in Bioengineering and Biotechnology 12, 1428189 (2024).

[35] Mallet, L. et al. Stimulation of subterritories of the subthalamic nucleus reveals its role in the integration of the emotional and motor aspects of behavior. Proceedings of the National Academy of Sciences 104, 10661–10666 (2007).

[36] Lang, P. J., Bradley, M. M. & Cuthbert, B. N. International affective picture system (1988).

[37] Belin, P., Fillion-Bilodeau, S. & Gosselin, F. The montreal affective voices: A validated set of nonverbal affect bursts for research on auditory affective processing. Behavior research methods 40, 531–539 (2008).

[38] Nastase, S. A., Goldstein, A. & Hasson, U. Keep it real: rethinking the primacy of experimental control in cognitive neuroscience. NeuroImage 222, 117254 (2020).

[39] Matusz, P. J., Dikker, S., Huth, A. G. & Perrodin, C. Are we ready for real-world neuroscience? (2019).

[40] Zaki, J. & Ochsner, K. N. The neuroscience of empathy: progress, pitfalls and promise. Nature neuroscience 15, 675–680 (2012).

[41] Falciglia, S. et al. Transformer-based long-term predictor of subthalamic beta activity in parkinson’s disease. npj Parkinson’s Disease 11, 210 (2025).

[42] Sonkusare, S., Breakspear, M. & Guo, C. Naturalistic stimuli in neuroscience: critically acclaimed. Trends in cognitive sciences 23, 699–714 (2019).

[43] Zacks, J. M., Speer, N. K., Swallow, K. M., Braver, T. S. & Reynolds, J. R. Event perception: a mind-brain perspective. Psychological bulletin 133, 273 (2007).

[44] Siegel, M., Donner, T. H. & Engel, A. K. Spectral fingerprints of large-scale neuronal interactions. Nature Reviews Neuroscience 13, 121–134 (2012).

[45] Brittain, J.-S. & Brown, P. Oscillations and the basal ganglia: motor control and beyond. Neuroimage 85, 637–647 (2014).

[46] Mathiopoulou, V. et al. Modulation of subthalamic beta oscillations by movement, dopamine, and deep brain stimulation in parkinson’s disease. npj Parkinson’s Disease 10, 77 (2024).

[47] Zavala, B. A., Jang, A. I. & Zaghloul, K. A. Human subthalamic nucleus activity during non-motor decision making. Elife 6, e31007 (2017).

[48] Leventhal, D. K. et al. Basal ganglia beta oscillations accompany cue utilization. Neuron 73, 523–536 (2012).

[49] Averna, A., Marceglia, S., Priori, A. & Foffani, G. Amplitude and frequency modulation of subthalamic beta oscillations jointly encode the dopaminergic state in parkinson’s disease. npj Parkinson’s Disease 8, 131 (2022).

[50] Swinnen, B. E. et al. Pitfalls and practical suggestions for using local field potential recordings in dbs clinical practice and research. Journal of Neural Engineering 22, 014001 (2025).

[51] Isaias, I. U. et al. Chronic adaptive versus conventional deep brain stimulation in parkinson’s disease: a blinded randomized pilot trial. medRxiv 2025–02 (2025).

[52] Arlotti, M. et al. A new implantable closed-loop clinical neural interface: first application in parkinson’s disease. Frontiers in neuroscience 15, 763235 (2021).

[53] Marceglia, S. et al. Double-blind cross-over pilot trial protocol to evaluate the safety and preliminary efficacy of long-term adaptive deep brain stimulation in patients with parkinson’s disease. BMJ open 12, e049955 (2022).

[54] Bentaleb, A. et al. Toward one-second latency: Evolution of live media streaming. IEEE Communications Surveys & Tutorials (2025).

[55] Bouten, C. V., Westerterp, K., Verduin, M. & Janssen, J. Assessment of energy expenditure for physical activity using a triax—ai. Age 23, 21–27 (1994).

[56] Bouten, C. V., Koekkoek, K. T., Verduin, M., Kodde, R. & Janssen, J. D. A triaxial accelerometer and portable data processing unit for the assessment of daily physical activity. IEEE transactions on biomedical engineering 44, 136–147 (1997).

[57] Horn, A. & Kühn, A. A. Lead-dbs: a toolbox for deep brain stimulation electrode localizations and visualizations. Neuroimage 107, 127–135 (2015).

