## Extended Data and Supplementary Materials for "Locked to the match: subthalamic engagement in sport match viewing"

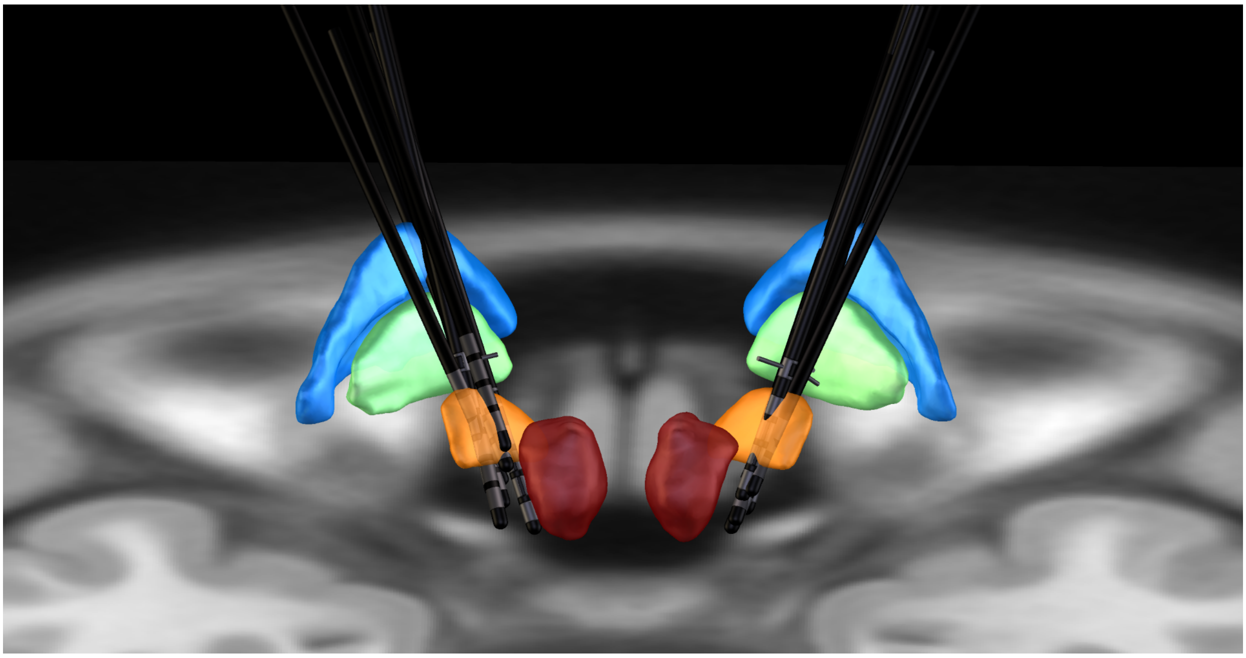

**Fig. Extended Data 1**: **3D localization of DBS leads.** Representative 3D reconstruction of deep brain stimulation (DBS) electrode placement within the STN for the patient cohort. Anatomical structures are color-coded as follows: red nucleus (red), STN (orange), internal globus pallidus (GPi, green), and external globus pallidus (GPe, blue). Electrode trajectories and contact positions were localized utilizing pre- and post-operative neuroimaging with the Lead-DBS toolbox[57].

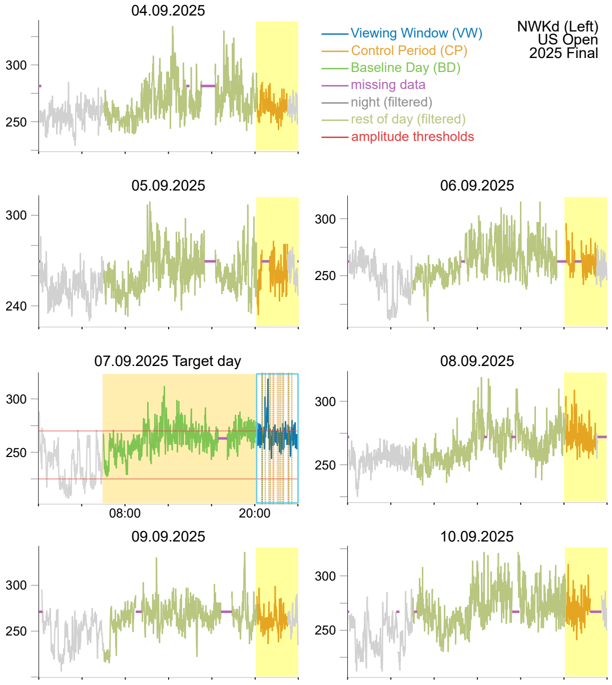


**Fig. Extended Data 2**: **Example of full monitoring period overview.** Left STN P-range amplitude time-series (1-min resolution) are shown for the entire monitoring period relative to a single live sport match. The aDBS amplitude threshold parameters for the driver channel are indicated by red horizontal lines on the target day. Missing data from the aDBS device, primarily due to charging interruptions, are marked by violet lines. Data from the left STN of patient NWKd while watching the US Open 2025 Final are visualized.

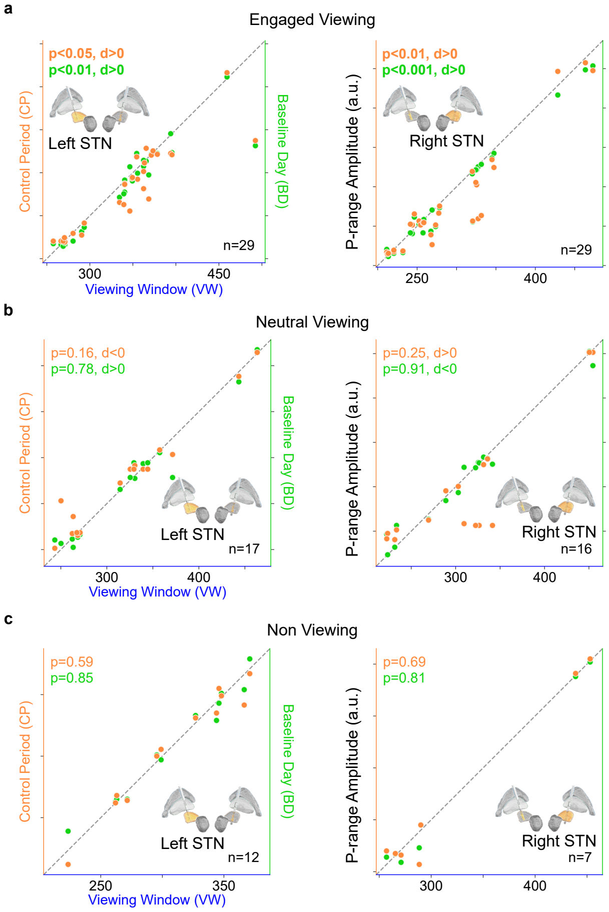


**Fig. Extended Data 3**: **Engagement effect over viewing and non-viewing categories.** Wilcoxon signed-rank tests (paired, non-parametric) were applied to median pairs for VW-CP and VW-BD for the three categories of engaged VWs (n=29 viewings, bilateral, 6 patients), neutral VWs (n=17 viewings, bilateral, 8 patients), and non-viewing (n=12 viewings left STN, n=7 viewings right STN, 6 patients). Significant positive effects ($p < 0.05$) were observed in both channels for both references only within the engaged VWs group, while no significant effects were found in the neutral and non-viewing categories. Population-level analysis results were confirmed through linear mixed effects modeling. Results are shown in scatter plot format with diagonal line of equality, with two y-axes for CP and BD P-range amplitude values and one x-axis for VW P-range amplitude values.

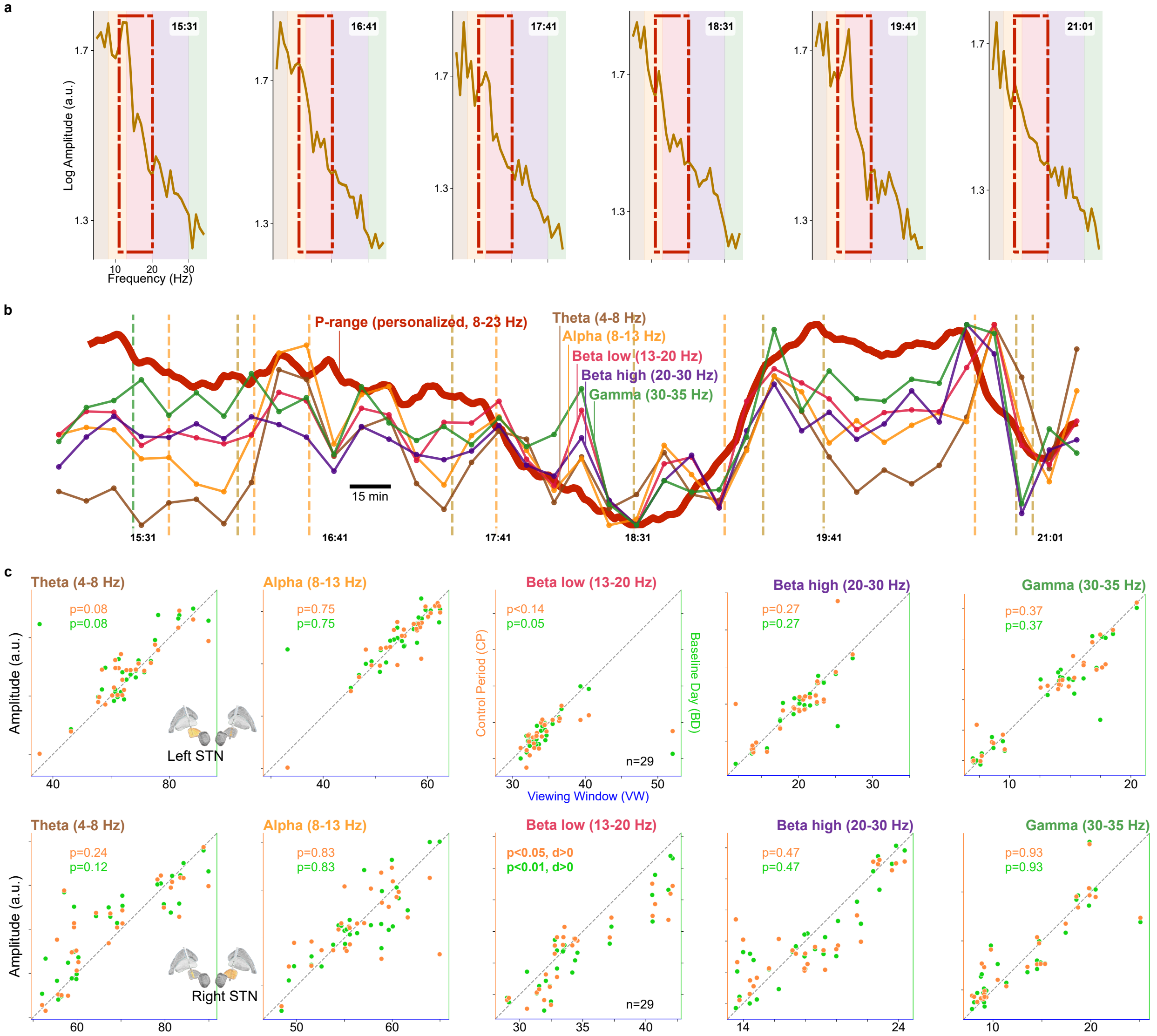


**Fig. Extended Data 4**: **Engagement effect over frequency bands. a,** Examples of amplitude spectra extracted by the AlphaDBS device. **b,** P-range trend and classic frequency bands amplitude profiles for one example VW (patient NWKb left STN watching the Roland Garros 2025 Final). **c,** Scatter plots (with diagonal line of equality) showing population-level results for each classic frequency band (Wilcoxon signed-rank tests with FDR correction). In addition to the 1-min resolution STN-LFPs amplitude in the P-range, the AlphaDBS device (Newronika SpA) returns directly the 10-min resolution amplitude spectra in the 5-34 Hz range. This allowed us to perform population-level analysis also for each classic frequency band, with median values now resulting from 10-min resolution time-series. All amplitude values are expressed in arbitrary units (a.u.). Specifically: (i) amplitude spectra are shown on log10 scale for visualization, (ii) 1-min resolution P-range amplitude is normalized to total amplitude spectrum since these are the data on which the final stimulation current is calibrated, (iii) 10-min resolution classic frequency band amplitude is computed as the mean spectral amplitude without normalization to total spectrum.

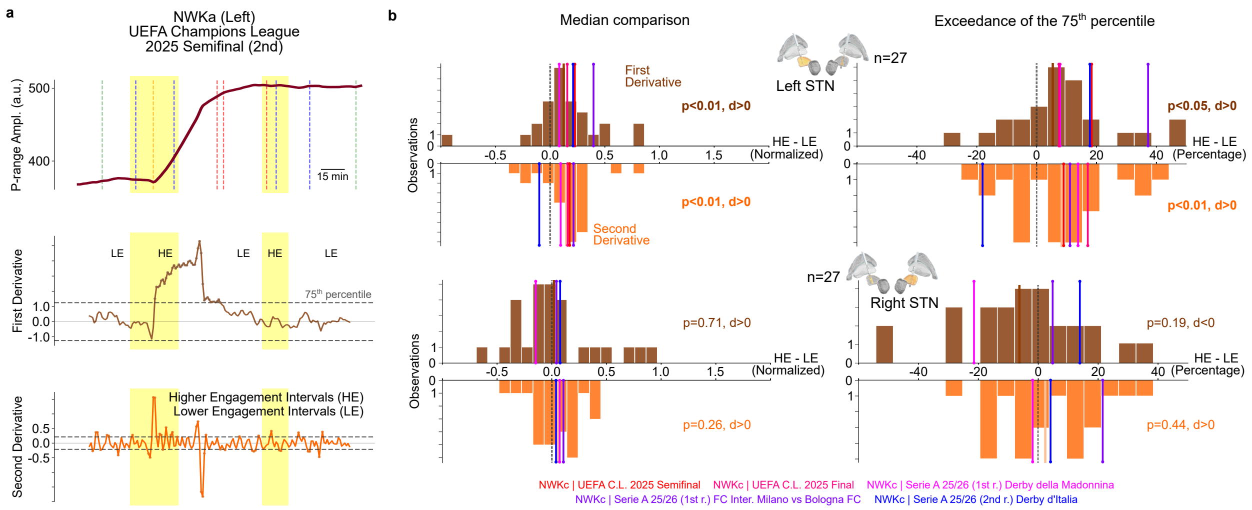


**Fig. Extended Data 5**: **STN P-range trend second derivative. a,** Engaged VWs trend's derivatives were estimated through the Savitzky-Golay Filter and evaluated through pairs of median values (HE, LE) extracted for each VW computing their absolute-valued magnitude and the percentage of time points exceeding their $75^{th}$ percentile threshold. First and second derivatives of the left STN P-range trend from patient NWKa while watching UEFA Champions League 2025 Semifinal (2nd leg) are visualized. **b,** Wilcoxon signed-rank tests (paired, non-parametric) were applied to median pairs for HE-LE for the group of engaged VWs (n=13). Significant positive effects ($p < 0.05$) were observed for both derivatives only in the left hemisphere, evaluating both absolute-valued magnitude and percentage of time points exceeding threshold. Population-level analysis results are shown in histogram format with null-line of equality showing the normalized difference HE-LE for absolute-valued magnitude and the difference in percentage for the threshold exceedance.

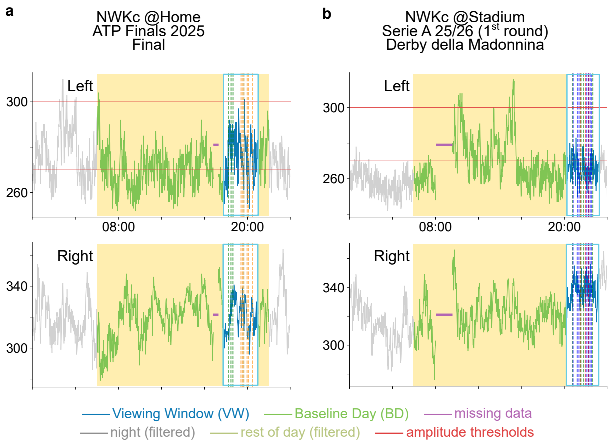


**Fig. Extended Data 6**: **Home viewing vs Stadium viewing target days. a,** Patient NWKc watching the ATP Finals 2025 Final, home viewing. **b,** Patient NWKc watching the Serie A 2025/2026 (1st round) Derby della Madonnina, stadium viewing. According to post-viewing interview, the patient left home around 5:00 pm to go to the stadium, highlighting differences in the neural signal between the two conditions at home (before) and outside in a higher motor-demanding condition (driving, taking transportations, walking, standing).

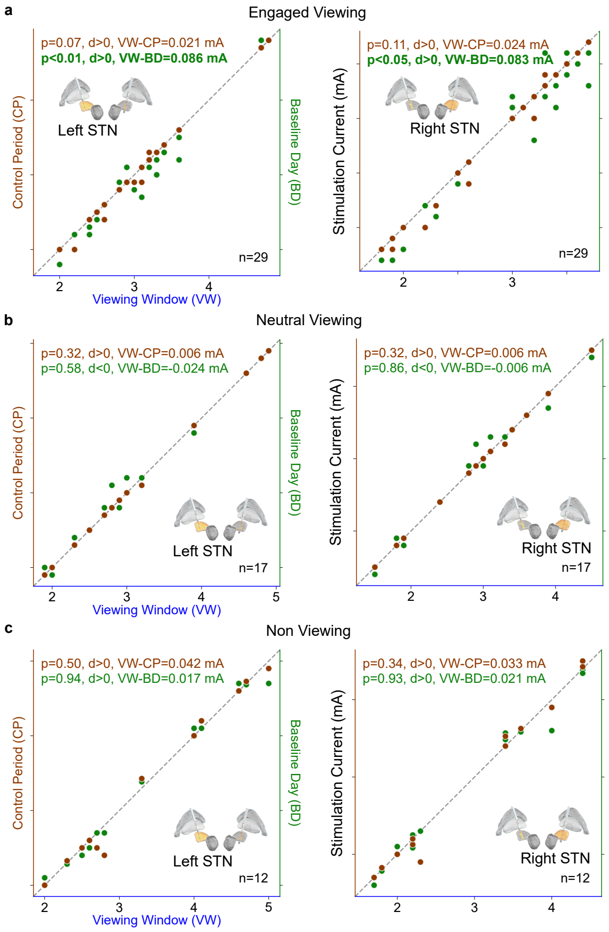


**Fig. Extended Data 7**: **Stimulation current across engagement levels.** The AlphaDBS device (Newronika SpA) records the history of bilateral stimulation current delivered to the STN through a linear proportional algorithm presenting two non-linearities at the minimum and maximum stimulation values. Population-level analysis of bilateral stimulation current (Wilcoxon signed-rank tests) revealed significantly higher stimulation during engaged VW compared to references CP and BD. The magnitude of stimulation increase ($\approx 0.12mA$) was modest but consistent across both hemispheres.

**Table Extended Data 1**: Clinical characteristics.

| Patient | Match | Monitoring period | Driver Channel | Stimulation Frequency (Hz) | Personalized Frequency range (P-range, Hz) | Power thresholds (a.u.) | Min-max stimulation currents (mA) | Drugs Therapy (LEDD) |
|---|---|---|---|---|---|---|---|---|
| NWKa | CLsf 2025 | 01/05/2025 - 09/05/2025 | Ch2 (Rx) | 130 | Ch1: [10, 20]<br>Ch2: [10, 20] | 410-470 | Ch1: [2.3, 3.0]<br>Ch2: [1.6, 2.3] | 400 |
| | | 09/05/2025 - 12/05/2025 | | 180 | | | Ch1: [2.0, 2.5]<br>Ch2: [1.4, 2.0] | |
| | CLf 2025 | 24/05/2025 - 04/06/2025 | | | | | | 450 |
| | | 04/06/2025 - 07/06/2025 | | | | | Ch1: [2.0, 2.7]<br>Ch2: [1.4, 2.0] | |
| | RGf 2025 | 01/06/2025 - 04/06/2025 | | | | | Ch1: [2.0, 2.5]<br>Ch2: [1.4, 2.0] | |
| | | 04/06/2025 - 12/06/2025 | | | | | Ch1: [2.0, 2.7]<br>Ch2: [1.4, 2.0] | |
| | | 12/06/2025 - 15/06/2025 | | | | | | 300 |
| | Wf 2025 | 06/07/2025 - 20/07/2025 | | | | | | 450 |
| | USOf 2025 | 01/09/2025 - 02/09/2025 | | 130 | | | | 600 |
| | | 02/09/2025 - 13/09/2025 | | 180 | | | Ch1: [1.7, 2.4]<br>Ch2: [1.2, 1.8] | |
| | ATPF 2025 | 01/11/2025 - 19/11/2025 | | | | 380-435 | Ch1: [1.9, 2.5]<br>Ch2: [1.4, 1.9] | |
| NWKb | CLf 2025 | 24/05/2025 - 04/06/2025 | Ch1 (Lx) | 130 | Ch1: [11, 20]<br>Ch2: [14, 23] | 320-360 | Ch1: [2.6, 3.6]<br>Ch2: [2.7, 3.7] | 350 |
| | | 04/06/2025 - 07/06/2025 | | | | | | |
| | RGf 2025 | 04/06/2025 - 11/06/2025 | | | | | | |
| | Wf 2025 | 29/06/2025 - 16/07/2025 | | | | | | |
| | USOf 2025 | 01/09/2025 - 07/09/2025 | | | | | | |
| | ATPF 2025 | 01/11/2025 - 20/11/2025 | | | Ch1: [11, 20]<br>Ch2: [11, 20] | | | |

*Continued on next page*

Clinical characteristics. (continued)

| **Patient** | **Match** | **Monitoring period** | **Driver Channel** | **Stimulation Frequency (Hz)** | **Personalized Frequency range (P-range, Hz)** | **Power thresholds (a.u.)** | **Min-max stimulation currents (mA)** | **Drugs Therapy (LEDD)** |
|---|---|---|---|---|---|---|---|---|
| | AO 2026<br>Coppa Italia<br>SerA Com-Ata | 25/01/2026 - 03/02/2026 | | | Ch1: [5, 10]<br>Ch2: [13, 18] | 335-395 | | |
| | SerA Com-Fio | 12/02/2026 - 15/02/2026 | | | | 310-395 | Ch1: [2.8, 3.6]<br>Ch2: [2.9, 3.7] | |
| NWKc | CLsf 2025 | 01/05/2025 - 12/05/2025 | Ch1 (Lx) | 130 | Ch1: [12, 17] | 200-280 | Ch1: [1.8, 2.6]<br>Ch2: [1.8, 2.6] | 825 |
| | CLf 2025 | 24/05/2025 - 07/06/2025 | | | | | | 875 |
| | RGf 2025 | 01/06/2025 - 15/06/2025 | | | Ch1: [11, 17]<br>Ch2: [11, 17] | | | |
| | USOf 2025 | 01/09/2025 - 15/09/2025 | | | | | | |
| | ATPF 2025 | 07/11/2025 - 19/11/2025 | | | | 270-300 | | 425 |
| | SerA Int-Mil | 17/11/2025 - 23/11/2025 | | | | | | |
| | SerA Int-Bola | 02/01/2026 - 07/01/2026 | | | | 255-300 | | 475 |
| | | 07/01/2026 - 09/01/2026 | | | | 290-340 | | |
| | AO 2026<br>SerA Cre-Int | 28/01/2026 - 02/02/2026 | | | | 250-335 | Ch1: [1.9, 2.6]<br>Ch2: [1.8, 2.6] | |
| | SerA Int-Juv | 13/02/2026 - 16/02/2026 | Ch2 (Rx) | | | 290-370 | | 635 |
| NWKd | CLsf 2025 | 29/04/2025 - 13/05/2025 | Ch1 (Lx) | 130 | Ch1: [11, 16]<br>Ch2: [11, 16] | 230-275 | Ch1: [3.8, 5.0]<br>Ch2: [2.2, 4.0] | 0 |
| | CLf 2025 | 24/05/2025 - 07/06/2025 | | | | | | |
| | RGf 2025 | 01/06/2025 - 15/06/2025 | | | | | | |
| | Wf 2025 | 06/07/2025 - 08/07/2025 | | | | | | |
| | | 08/07/2025 - 16/07/2025 | | | | | | |
| | | 16/07/2025 - 20/07/2025 | | | | | Ch1: [4.0, 5.0]<br>Ch2: [2.2, 4.0] | |

*Continued on next page*

Clinical characteristics. (continued)

| Patient | Match | Monitoring period | Driver Channel | Stimulation Frequency (Hz) | Personalized Frequency range (P-range, Hz) | Power thresholds (a.u.) | Min-max stimulation currents (mA) | Drugs Therapy (LEDD) |
|---|---|---|---|---|---|---|---|---|
| | USOf 2025 | 01/09/2025 - 08/09/2025 | | | | 225-270 | Ch1: [4.3, 5.0]<br>Ch2: [2.4, 4.0] | 100 |
| | | 08/09/2025 - 12/09/2025 | | | | 230-285 | Ch1: [4.0, 5.0]<br>Ch2: [2.2, 4.0] | |
| | | 12/09/2025 - 14/09/2025 | | | | 225-280 | Ch1: [4.3, 5.0]<br>Ch2: [2.4, 4.0] | |
| | ATPF 2025 | 01/11/2025 - 17/11/2025 | | | | | | 0 |
| | SerA Juv-Nap<br>AO 2026<br>SerA Par-Juv | 24/01/2026 - 02/02/2026 | | | | | | |
| | SerA Int-Juv | 13/02/2026 - 16/02/2026 | | | | | | |
| NWKe | CLsf 2025 | 29/04/2025 - 13/05/2025 | Ch2 (Rx) | 130 | Ch1: [11, 20]<br>Ch2: [8, 11] | 230-290 | Ch1: [2.2, 3.0]<br>Ch2: [1.7, 2.6] | 350 |
| | CLf 2025 | 24/05/2025 - 07/06/2025 | | | | | | 450 |
| | RGf 2025 | 01/06/2025 - 15/06/2025 | | | | | | |
| | Wf 2025 | 06/07/2025 - 20/07/2025 | | | | | | |
| | USOf 2025 | 01/09/2025 - 15/09/2025 | | | | 240-300 | Ch1: [2.3, 3.0]<br>Ch2: [1.8, 2.6] | 350 |
| | ATPF 2025 | 13/11/2025 - 20/11/2025 | | | | | | |
| NWKf | CLsf 2025 | 29/04/2025 - 08/05/2025 | Ch1 (Lx) | 90 | Ch1: [11, 17]<br>Ch2: [11, 17] | 275-335 | Ch1: [3.8, 4.6]<br>Ch2: [4.2, 4.8] | 1167.5 |
| | Wf 2025 | 06/07/2025 - 20/07/2025 | | | | | | |
| | USOf 2025 | 02/09/2025 - 11/09/2025 | | | | 235-300 | Ch1: [3.5, 4.3]<br>Ch2: [4.2, 4.8] | 1001.25 |
| NWKg | ATPF 2025 | 01/11/2025 - 14/11/2025 | Ch2 (Rx) | 130 | Ch1: [9, 15]<br>Ch2: [12, 18] | 220-275 | Ch1: [2.4, 2.9]<br>Ch2: [2.7, 3.1] | 850 |
| | | 14/11/2025 - 17/11/2025 | | | | 210-250 | | |
| | AO 2026<br>SerA Cre-Int | 29/01/2026 - 02/02/2026 | | | | 200-260 | | |

*Continued on next page*

Clinical characteristics. (continued)

| **Patient** | **Match** | **Monitoring period** | **Driver Channel** | **Stimulation Frequency (Hz)** | **Personalized Frequency range (P-range, Hz)** | **Power thresholds (a.u.)** | **Min-max stimulation currents (mA)** | **Drugs Therapy (LEDD)** |
|---|---|---|---|---|---|---|---|---|
| | SerA Int-Juv | 13/02/2026 - 16/02/2026 | | | | 210-280 | | |
| NWKh | ATPF 2025 | 01/11/2025 - 19/11/2025 | Ch1 (Lx) | 130 | Ch1: [12, 18] | 210-300 | Ch1: [1.7, 2.5]<br>Ch2: [1.7, 2.3] | 450 |

*End Table*

**Table Extended Data 2**: Patients monitored across our DBS studies.

| Patients → Our studies ↓ | n.1 | n.2 | n.3 | n.4 | n.5 | n.6 | n.7 | n.8 | n.9 | n.10 | n.11 | n.12 |
|---|---|---|---|---|---|---|---|---|---|---|---|---|
| **[17]Caffi et al. 2024** | Pt | | | | | | | | | | | |
| **[34]Isaias, Caffi et al. 2024** | | Pt | | | | | | | | | | |
| **[51]Isaias, Marceglia et al. 2025** | Pt.5 | Pt.7 | Pt.8 | Pt.11 | Pt.12 | | | Pt.15 | | | | |
| **[41]Falciglia et al. 2025** | NWK2 | | NWK3 | | NWK1 | | | NWK4 | | | | |
| **Falciglia et al. 2026** | NWKd | | | NWKf | NWKb | | NWKa | | NWKh | NWKg | NWKc | NWKe |

**Table Extended Data 3**: Post-viewing interview protocol.

| **Question Type** | **Content** |
|---|---|
| **Opening question (free response)** | Did you watch the match? Did you feel engaged while watching it? Please speak freely about that experience. *[Hai visto la partita? Ti sei sentito coinvolto nel guardarla? Parlami liberamente di quel momento.]* |
| **Structured questions** | **1. Viewing context.** Where did you watch the match (home [TV channel, sitting, lying down] or stadium)? Did you join after the start or turn off/switch channels before the end? If so, when? *[Dove hai visto la partita (casa [canale TV, seduto, sdraiato] o stadio)? Ti sei collegato dopo l'inizio o hai spento o cambiato canale prima della fine? Quando?]*<br>**2. Emotional dynamics (i).** How did you feel and what emotions did you experience during each set/period of the match? *[Come stavi e cosa provavi durante ogni set/tempo della partita?]*<br>**3. Emotional dynamics (ii).** Did you experience anger, sadness, confidence, or lapses in attention during the match? *[Hai provato rabbia, tristezza, fiducia, o cali di attenzione durante la partita?]*<br>**4. Cognitive appraisal: outcome expectation and resignation.** Did you ever think the match would end with a certain result? When did you resign yourself to the outcome? *[Hai mai pensato la partita potesse finire con un certo risultato? Quando ti sei rassegnato al risultato?]*<br>**5. Comparative engagement.** Compared to previous matches, did this one engage you more, less, or equally? In this match, did you feel more disappointed, dejected, or experience other emotions? *[Rispetto alle passate partite, questa ti ha coinvolto di più, di meno o uguale? In questa ti sei sentito più deluso o più giù o altro?]* |

### Supplementary Materials for:

### Locked to the match: subthalamic engagement in sport match viewing

Salvatore Falciglia[1,2], Laura Caffi[1,2,3,4], Fabrizio Luiso[3,4], Chiara Palmisano[3], Alberto Mazzoni[1,2†], Ioannis Ugo Isaias[3,4†]

[1]The BioRobotics Institute, Scuola Superiore Sant'Anna, 56025, PI, Italy
[2]Department of Excellence in Robotics and AI, Scuola Superiore Sant'Anna, 56025, PI, Italy
[3]University Hospital Wuerzburg and Julius Maximilian University of Wuerzburg, 97080, Wuerzburg, Germany
[4]Parkinson Institute Milan, ASST G. Pini-CTO, 20126, MI, Italy

[†]These authors gave equal senior contribution

**Table Supplementary Information 1**: Demographic characteristics.

| Patient | Age (y) | Disease duration (y) | Time since surgery (y) | Lead | Handed -ness | Side of onset | Clinical phenotype | MDS-UPDRS III* (STIM-ON MEDS-ON) |
|---|---|---|---|---|---|---|---|---|
| NWKa | 55-65 | 10-15 | 0.5 | Abbott 6172 | Right | Right | Tremor dominant | 17 |
| NWKb | 45-55 | 15-20 | 3 | Medtronic 3389 | Right | Left | Akinetic-rigid | 5 |
| NWKc | 55-65 | 10-15 | 1 | Abbott 6172 | Right | Right | Akinetic-rigid | 9 |
| NWKd | 45-55 | 15-20 | 8 | Medtronic 3389 | Right | Right | Akinetic-rigid | 13** |
| NWKe | 55-65 | 20-25 | 2 | Medtronic 3389 | Right | Right | Akinetic-rigid | 14 |
| NWKf | 55-65 | 15-20 | 7 | Medtronic 3389 | Right | Right | Akinetic-rigid | 30 |
| NWKg | 65-75 | 15-20 | 1.5 | Abbott 6172 | Right | Right | Akinetic-rigid | 9 |
| NWKh | 65-75 | 15-20 | 1.5 | Abbott 6172 | Right | Right | Akinetic-rigid | 10 |

*: data collected for all patients in July 2025
**: the patient does not take dopaminergic therapy

**Table Supplementary Information 2**: Post-viewing interview transcriptions.

| | | |
|---|---|---|
| Engaged Viewings | NWKa UEFA Champions League 2025 Semifinal (2nd leg) | ”I was feeling confident — I really thought Inter would win, both before and after the 2–2 draw [...] Then, when it went to 3–2, I started thinking, okay, maybe we’ve actually lost this one [...] But when Acerbi scored, right at the very end, I had to believe again [...] My focus stayed sharp the whole game, from start to finish.” |
| | NWKa UEFA Champions League 2025 Final | ”There was this slow build-up of anger — not really because of the match itself, but because of my neighbor (a Juventus fan) [...] Already at 3–0 I’d written it off as lost, then at 4–0 it was hopeless — I switched the channel [...] When I changed the channel [the interviewer asks: ‘did you get distracted?’] well, yes and no — it’s just that my neighbor kept bugging me the whole time.” |
| | NWKa ATP Finals 2025 Final | ”I was able to watch up until 4-4. I remember the final rallies (before 4-4) more vividly [...] and the interruption during the first set (due to a spectator’s medical emergency) [...]” |
| | NWKb Roland Garros 2025 Final | ”By the third set I was relaxed — for me, he (Sinner) had it in the bag [...] But then, when he had those three match points in the fourth set, I couldn’t even sit down. I actually turned off the TV for ten minutes, convinced he was gonna lose [...] Then I turn it back on — and he’d broken back! I thought, that’s it, we’ve got this! But no... turns out, we didn’t.” |
| | NWKb Wimbledon 2025 Final | ”That one was less emotional for me [...] I mean, I really thought he had it again. He lost the first set, but I wasn’t too bothered [...] Then after he took the second and leveled it up, that was it — I was completely sure he’d win.” |
| | NWKb US Open 2025 Final | ”The first set just slipped away — gone. I didn’t feel much, stayed pretty neutral [...] Then in the second set, when he finally managed to break, that’s when I felt real joy — no anxiety at all [...] During the third set there wasn’t anxiety either, more like surprise, amazement really. But I still believed. Up until the fourth set, when it was 2–2, before Alcaraz broke him [...] And then, well... resignation kicked in during those last four games of the final set.” |
| | NWKb ATP Finals 2025 Musetti - De Minaur | ”[...] I watched the match in an engaged way [...] Then at the end during the celebration, (Musetti) went absolutely crazy [the patient laughs] [...] He was right to celebrate like that, he played a beautiful match, it’s just that we’ve been spoiled (by Sinner)” |
| | NWKb ATP Finals 2025 Sinner - De Minaur | ”Well, I’d say that in recent days (Sinner) had been hiding a bit, but today what a match! [...] The point with which he won the first set was definitely spectacular” |
| | NWKb ATP Finals 2025 Final | ”I saw Sinner’s splendid victory! I made a list of the moments where I was most thrilled [...] Until the end it could have gone either way, very balanced [...] Hard to stay calm [...] Let’s say I watched almost all of it standing up [...]” |

*Continued on next page*

| | |
|---|---|
| NWKb Coppa Italia 2025/2026 Fiorentina - Como | ”We conceded a goal immediately and then we equalized with a bit of luck. [...] Fiorentina at the beginning was also playing well, they were pressing us high. [...] But immediately after there was no match anymore [...] Douvikas got injured and Nico Paz came on, he hadn’t started last night [...] They scored after this goal, there was a whole scramble [...] In the second half there was no match anymore [...] The substitutions struck me [...] Nico Paz came on, then Perrone, Addai who came back [...] and then the whole team kinda (struck me) we were playing with reserves [...] All quick play, not holding the ball, bam they play from memory, all one-touch passes [...] Nico Paz scored a beautiful goal yes but the goalkeeper also made a bit of a blunder [...] Then Fiorentina didn’t take another shot [...] Later he took off Nico Paz, brought on Morata who got unblocked. He scored a beautiful goal [...] Kuhn beautiful pass, Morata was clearly onside and scored. [...] Between the first and second half they finished setting up (the polysomnography equipment) and I watched the second half alone calmly sitting in bed (in hospital admitted for the ongoing study). The first half I was watching the match commenting on it with the engineer and the doctor while the equipment was being set up (sitting raising my arms or moving my body to facilitate the equipment setup) [...] Never anxious, I was sure we would win even when we were a goal down. I was confident [...]” |
| NWKb Australian Open (Semif.) Alcaraz - Zverev | ”I started watching it from the beginning. I was getting angry, really annoyed thinking that Zverev always has to get his points stolen and get broken even when he seems to play well [...] Bah he does it on purpose [...] So around 7:00 with Alcaraz up 2-0 I went to bed. Then I turned it back on around 8:00 and saw they were still playing, actually they were at the tie-break that Zverev won. [...] The third and fourth set Zverev won [...] At one point I thought Zverev could win the match [...] The whole fifth set was played brilliantly by both. Zverev had even broken him, but then you could see Zverev was completely drained in fact then (Alcaraz) broke him back. [...] It finished around 10:30, even after [...] The fourth set tie-break was also played very well by both [...]” |
| NWKb Australian Open (Semif.) Sinner - Djokovic | ”I was truly embittered, come on you can’t lose like that. [...] Sinner’s first set was beastly, extremely high level. Then instead he got broken immediately. I thought he’d manage to break back and instead [...] From the third set, even though he’d broken him, and then he won but he was already declining. In the fifth set then he wasn’t Sinner anymore, he had 18 break points between fourth and fifth set, but absolutely crazy, he didn’t convert even one. Djokovic instead all the ones he had, very few, he converted them all. And anyway Sinner had 26 aces an absolute record. He also had more than 60 winners against Djokovic’s 48-49, yet he lost. [...] On the saved match points, the second was insane. There I made a racket, I even made some rude gestures... (in hindsight) I’m not very sporting I think [...] In the third set I had to open the door for the mailwoman, then my son, then messages from my wife, but I didn’t have lunch, I was watching the match [...]” |

*Continued on next page*

| | |
|---|---|
| NWKb Australian Open 2025 Final | ”I didn’t like the final! [...] I started watching from the end of the first set [...] The only semblance of a match was the end of the fourth set when there was a bit of suspense [...] Djokovic played well but after a while he couldn’t manage physically anymore [...] They barely reached 3 hours of match [...] The only nice moment is when Djokovic tried to take it to the fifth. He had the break point, just like with Sinner, but Alcaraz didn’t give him a chance [...] I had lunch after the match [...]” |
| NWKb Serie A 2025/2026 Como - Atalanta | ”I saw Como, they played a good match eh [...] Atalanta’s goalkeeper best on the pitch saved everything. [...] I wasn’t expecting the penalty [...] Then when I saw that only Nico Paz was left to take it I didn’t even watch, I knew he’d miss it [...] And despite it being over, we still managed to have a huge opportunity in those last 30 seconds [...] They played a stoic match, our goalkeeper only made one save [...] The slap in the first half that led to the red card, there I said – It’s done! – and instead no [...] At least seven, eight clear chances we had, Nico Paz had two chances, Duvikas wasted a goal already made he kicked it at two miles per hour. [...] First half Como played well. The second half Atalanta had some very nice counterattacks but they didn’t even get to shoot [...]” |
| NWKb Serie A 2025/2026 Como - Fiorentina | ”I saw the match alone, sitting on the couch, I got up a few times for the match but you have everything in the sensors [...] The final result is quite deceptive because it could have ended even worse. He took off all the defenders, and then Morata got himself sent off [...] First half so-so, you could see Fiorentina was spreading out in fact then they scored on us. Nico Paz unwatchable, Baturina instead in great form, but alone... [...] We gifted them the first half [...] Second half, after 10 min stupidity and immediately penalty for Fiorentina, there was still more than half an hour left [...] He brought on Morata. After which we pushed, but nothing, he put in all offensive people and we found a goal, actually an own goal, when there was a quarter hour left. Then there were some provocations, Morata fell for it and got himself sent off [...] It was an ugly match, but ugly, really pitiful [...] After 10 min I said ’Look, today we’re losing!’. I was a bit resigned [...] I celebrated at Como’s goal, maybe luckily we would have then found a second goal, but really nothing nothing [...]” |
| NWKc US Open 2025 Final | ”[...] I watched the match but I was dozing off at times. Between the third and fourth set especially I fell asleep [...] When Alcaraz won the first set I wasn’t worried, Sinner never gives up. Until the second I was actually still confident (in Sinner’s victory). At the third set I was hoping for a fourth and fifth set. Then in the fourth set resignation set in [...] In the final minutes, when Sinner canceled Alcaraz’s two match points, that’s when I started hoping a bit” |
| NWKc ATP Finals 2025 Sinner - Zverev | ”I watched the match in an engaged way, it didn’t last long! [The patient laughs] Sinner played very well [...]” |
| NWKc ATP Finals 2025 Final | ”[...] The moments that moved me most were the tie-break and the last game of the second set” |
| NWKc Australian Open (Semif.) Alcaraz - Zverev | ”From 9 I saw Alcaraz’s match, they were in the fifth (set) [...] The set was quite hard-fought [...] I thought Zverev would win, instead then Alcaraz never gives up [...] I couldn’t wait for it to finish to see the other (match) [...]” |

*Continued on next page*

| | |
|---|---|
| NWKc Australian Open (Semif.) Sinner - Djokovic | ”A bit of a disappointment [...] I dozed off a bit in the third set, I was so convinced Sinner would win. I woke up then in the fourth set [...] In the fifth there was a bit of emotion towards the end. You couldn’t see technical superiority anymore, it was more an endurance race [...] The only point where I got thrilled was when Sinner saved the second match point. It was a special point. I thought he’d get to the tie-break, but then the match point came [...] I had lunch around 12:45 until 1:15. I didn’t watch it there, I resumed watching after [...]” |
| NWKd ATP Finals 2025 Final | ”I’m not a fan but I watched the match in an engaged way [...] Let’s say I felt a fairly uniform calm until the tie-break of the first set where I had some jolts and...] (In the second set) the initial break made me jolt until the counter-break, when I recovered [...] The last rally was exciting” |
| NWKd<br>Serie A 2025/2026<br>Juventus - Napoli | ”I saw the whole match and it really struck me. Juve particularly faster, I experienced it in a more joyful way. [...] I remember and was really struck by the first goal. I truly enjoyed it, sportingly speaking [...] The 2-0 with Yildiz’s goal was just the pinnacle [...] I watched the match strictly alone [...]” |
| NWKd Australian Open (Semif.) Sinner - Djokovic | ”I really didn’t feel great emotions. As said, I’m not a big tennis fan even though I followed the whole match. [...] I connected around 8:50 – 9:00 and there was Alcaraz’s match which I also watched though with much less attention. With more attention I watched Sinner. [...] The first 3 games won by Sinner in the first set made me think of a downhill match. I wasn’t calm because I saw Djokovic more and more in the match especially from the second set. [...] My being a bit calmer or less depended in the end on the match score. It was very up and down. [...] One of the last points, at the first saved match point I thought, hoped, a bit for the miracle, but it wasn’t to be [...]” |
| NWKd<br>Serie A 2025/2026<br>Parma - Juventus | ”Beautiful emotions for Juventus, I started watching from 8:40 PM until shortly after the end [...] The break between first and second half lasted a bit longer [...] At Conceicao’s crossbar I jumped from my chair [...] Bremer’s first goal was a liberation [...] At the moment of Parma’s goal my arms dropped... Cambiaso’s own goal, the lowest point morale-wise [...] The 3-1 goal was also liberating because it sealed the victory. [...] At the fifth goal, then disallowed, I was already over the moon, it would have been excessive [...] As emotions, more joy. Before the goal a bit of anxiety, I saw Juventus playing well and I hoped they’d break through [...]” |
| NWKd<br>Serie A 2025/2026<br>Inter - Juventus | ”I saw the whole match at home alone [...] At the first Inter goal, actually Cambiaso’s own goal, I got a bit disheartened, but then I saw the match immediately turned good with Juventus’s second goal [...] Well then the whole match was a bit distorted by the double yellow on Kalulu [...] my arms dropped, total despair, for an invented expulsion by the referee [...] Also the rest of the match I watched it with Inter attacking more and more, so I watched it in a more subdued way [...] until then the 90th minute goal of Locatelli’s equalizer, I jumped from my chair [...] Then at the 93rd Inter scored eh well, total despair [...] Inter wasn’t doing who knows what but playing always in attack I took for granted that sooner or later the goal would come, and they did it, I recovered though right after at the equalizer goal and I said to myself ’So, there’s justice!’, and instead no there wasn’t” |

*Continued on next page*

| | | |
|---|---|---|
| | NWKe ATP Finals 2025 Final | ”I watched in an engaged way, more or less consistently throughout. [...] Mid-match Sinner was missing easy points. This struck me more than the points he made [...] At the end, the celebration on the ground was moving” |
| | NWKg ATP Finals 2025 Final | ”I watched until 7:30 PM [...] At certain moments mid-first set I was more convinced Alcaraz would win rather than Sinner.” [The patient was quite happy to watch the match at our request] |
| | NWKg<br>Serie A 2025/2026<br>Cremonese - Inter | ”I saw Inter yes, other goals could have gone in but okay be satisfied [...] First of all we had about 90% possession, there were no excessive worries [...] at the goal I was pleased, I didn’t celebrate who knows how, it wasn’t Inter-Milan or Inter-Juve. [...] In the second half Cremonese hit a post, an action that almost cost a goal. [...] Inter’s goals one more beautiful than the other. The first a header by Lautaro, the second Zielinski fired a rocket... the goalkeeper tried but it slipped from his hands, it tore through the net! [...]” |
| | NWKg<br>Serie A 2025/2026<br>Inter - Juventus | ”You’re seeing the disgrace the Juventus fans are making... when things were resolved in their favor everything was fine [...] now because one was sent off they’re taking it out on Inter [...] what do you want! [...] I saw the whole match, in company with a very dear friend of mine [...] When Inter went 2-1, when a certain Pio Esposito scored, we started saying loudly -Pio-pio-pio-pio!- we celebrated big time! [...] Then (at 3-2) even worse, the Polish guy’s blast, Zielinski, oh my, from outside the area, a strike at least 45 meters. [...] On Juve’s goals, honestly I was convinced that Inter one way or another would bring home the victory, because Inter’s game right now is really important compared to all the other teams [...] We watched the match sitting on the couch [...] All three of Inter’s goals were beautiful, in open play [...] The first goal was a shot by the Inter player badly deflected by a Juventus player and the goalkeeper, blameless, found the ball in the net, and similar was Juventus’s first goal [...] But there was still plenty of time, I didn’t experience it badly or worried [...] Juventus’s second goal came towards the end, so much so that then the Inter coach put in two strikers to win the match [...] On the red, I wasn’t happy, but the Juventus player shouldn’t have given the referee reason to think about a possible red [...]” |
| @Stadium Viewings | NWKc<br>UEFA Champions League 2025 Semifinal (2nd leg) | ”The match was very vibrant [...] At the beginning it was a crescendo because we went ahead, then when they equalized clearly a bit less [...] Then when we went behind I was a bit more dejected, we were almost convinced we were out [...] The strongest emotion was the goal at the 93rd minute, the equalizer, and then well also extra time [...] Then on the last goal it was maximum exhilaration” |
| | NWKc<br>UEFA Champions League 2025 Final | ”We left before the end, after 5-0, around the 88th minute. There were 3 of us. I’m always an optimist, so let’s say it’s rare for me to leave early, almost never, but one of the others wanted to go home after the third goal, and he was the one driving... We had a hard time keeping him there let’s say [...] There weren’t any moments of great excitement. It was a crescendo of depression [...] Until 2-0 I still believed, then let’s say we believed less [...] I didn’t feel anger or anxiety, more disappointment I’d say [...] Resignation came at 4-0” |

*Continued on next page*

| | | |
|---|---|---|
| | NWKc<br>Serie A 2025/2026<br>Derby della Madonnina | ”I left home at 5:30 PM and entered the stadium at 8:00 PM. [...] In the first half I believed, I saw both posts hit by Inter, especially the second was clearly visible. I felt a bit of anger. [...] After Milan’s goal I still believed, after the penalty I started believing less. [...] The coup de grâce was the missed penalty. [...] As time passed hope diminished” |
| | NWKc<br>Serie A 2025/2026<br>Inter - Bologna | ”I left home at 6:00 PM and arrived around 7:30 PM. There were 2 of us. We wandered around and then entered around 8:00 PM [...] There was the initial minute of silence for these days’ tragedy [...] Lively match. (Inter’s players) Played well but many wasted chances. [...] They finished the first half 1-0 but it could have been much more [...] anger about the result and the yellow cards [...] At a certain point chants even started (from the fans) against the referee [...] (In the second half) Incredible opportunity. Lautaro from 2 meters hit a spectacular crossbar. Then the goals scored, excellent! [...] The goal conceded 5 minutes from the end fairly inconsequential. [...] I experienced the second half more calmly. [...] Until the goal there was a bit of worry about those strange matches where maybe you have ten chances and then the opponent scores on their first attempt. [...] Not a worrying opponent but still a better match compared to the derby, also for the result let’s say!” |
| | NWKc<br>Serie A 2025/2026<br>Inter - Juventus | ”I saw the whole match at the stadium [...] I left around 5:00 PM, there were two of us, we entered around 8:00 PM [...] At the beginning the match was quite balanced, the best chances we’d perhaps had, we’d hit that double post, that was perhaps the most exciting moment [...] Then well there was the sending off that from there seemed correct, but well it wasn’t [...] Then obviously there were the goals (as most exciting moments), our first experienced with enthusiasm, the equalizer with disappointment [...] Then they were down to 10, you thought it would be easier instead we struggled to go ahead and then they equalized pretty quickly [...] For me the most important moment of the match was the third goal, by now we’d resigned ourselves to the draw [...] Most of the match I watched sitting, only at goals and the most important actions usually you stand up [...]” |
| Neutral Viewings | NWKa US Open 2025 Final | ”[...] I watched the tennis match sort of. I was much more interested in the final part of Italy’s basketball final [...] The tennis match didn’t excite me at all, barely anything [...]” |
| | NWKa ATP Finals 2025<br>Alcaraz - Musetti | ”Definitely a more interesting match than the national football team’s [...] at least until the end of the first set, then I fell asleep. [...] I channel-surfed between the tennis match and the football match.” |
| | NWKb ATP Finals 2025<br>Sinner - Zverev | ”I saw the match [...] Apart from a jolt for the drop shot that brought Sinner ahead by a set, I’d say everything was normal [...] It didn’t engage me much, you could see Sinner had control of the match” |
| | NWKb ATP Finals 2025<br>Alcaraz - Musetti | ”I saw the match [...] Unfortunately though the match didn’t offer that spectacle everyone was expecting [...]” |
| | NWKb ATP Finals 2025<br>Sinner - Shelton | ”If I have to tell the truth, (the match) didn’t excite me much [...] He (Sinner) was already qualified as first, the other (Shelton) was already eliminated [...]” |
| | NWKb ATP Finals 2025<br>Zverev - A.Aliassime | ”I saw the match only in small part [...] often changing channels [...] I don’t remember well which points (I watched)” |

*Continued on next page*

Post-viewing interview transcriptions. (continued)

| | |
|---|---|
| NWKb ATP Finals 2025 Alcaraz - A.Aliassime | ”I watched the match because it was a semifinal, but actually it didn’t excite me much [...]” |
| NWKc Australian Open 2025 Final | ”I only saw the fourth set, I connected around 12:00. [...] It interested me relatively, I was rooting for Djokovic [...] There wasn’t any strong emotion anyway [...]” |
| NWKc Serie A 2025/2026 Cremonese - Inter | ”The match was quite easy, the first goal came quite quickly [...] There weren’t great emotions, the firecracker that hit the goal-keeper struck me more [...] Cremonese then hit a post towards the end, there was a moment of suspense because the referee had given six minutes of stoppage time so it would have been special if they’d found a goal there [...]” |
| NWKd Wimbledon 2025 Final | ”[...] From the beginning I felt good about this one, I was confident Sinner would win. [...] I don’t remember anything else” |
| NWKd US Open 2025 Final | ”[...] I only saw the first set. I thought Sinner would lose the match. I fell asleep after the first set” |
| NWKd Australian Open 2025 Final | ”I saw from the second set until the end. [...] I’m not a huge fan but I liked it. I believed until the end that Djokovic would take it to the fifth set [...] No strong emotion anyway [...] I started having lunch towards the end of the match but I was still watching [...]” |
| NWKe Wimbledon 2025 Final | ”[...] It was an even match, I remember Sinner was playing well [...] Honestly I don’t remember anything else” |
| NWKe US Open 2025 Final | ”[...] I saw the first two sets, then I fell asleep. [...] I thought Alcaraz would win, he was playing so much better than Sinner, despite Sinner then winning the second set himself [...] I felt let’s say resignation, so much so that I was even telling my wife ’Today Sinner’s going to take a beating’ [...]” |
| NWKf US Open 2025 Final | ”[...] I saw the first half of the match, then I went to sleep. I had thought from the beginning that Sinner would lose [...]” |
| NWKg Australian Open (Semif.) Alcaraz - Zverev | ”I saw the match from 9:15 until the end [...] Look, tennis doesn’t excite me much [...] I wouldn’t even know how to identify who won or anything from the score they show on screen [...] In the end I seem to understand something from the players’ smiles and celebrations [...]” |
| NWKh ATP Finals 2025 Final | ”Yes I watched the match but it didn’t interest or excite me [...]” |

**Table Supplementary Information 3**: Per-viewing, per-channel comparisons across engagement levels.

| | Viewing | Ch1 (Lx) VW-CP p-value (Cohen's d) | Ch1 (Lx) VW-BD p-value (Cohen's d) | Ch2 (Rx) VW-CP p-value (Cohen's d) | Ch2 (Rx) VW-BD p-value (Cohen's d) |
|---|---|---|---|---|---|
| Engaged Viewings | NWKa UEFA Champions League 2025 Semifinal (2nd leg) | *** (1.43) | *** (2.27) | *** (1.67) | *** (1.06) |
| | NWKa UEFA Champions League 2025 Final | *** (0.76) | n.s. (-0.23) | * (0.16) | *** (1.20) |
| | NWKa ATP Finals 2025 Final | *** (0.78) | n.s. (0.02) | n.s. (0.02) | *** (0.72) |
| | NWKb Roland Garros 2025 Final | *** (0.70) | *** (0.53) | *** (1.48) | *** (1.52) |
| | NWKb Wimbledon 2025 Final | *** (0.38) | *** (0.61) | *** (1.40) | *** (1.35) |
| | NWKb US Open 2025 Final | *** (0.58) | *** (0.46) | *** (1.36) | *** (1.05) |
| | NWKb ATP Finals 2025 Musetti - De Minaur | * (0.12) | *** (0.50) | *** (1.68) | *** (1.50) |
| | NWKb ATP Finals 2025 Sinner - De Minaur | *** (0.41) | *** (0.51) | *** (1.51) | * (0.21) |
| | NWKb ATP Finals 2025 Final | *** (0.53) | n.s. (0.01) | *** (1.52) | *** (0.32) |
| | NWKc US Open 2025 Final | *** (1.14) | *** (0.87) | *** (1.49) | * (0.52) |
| | NWKc ATP Finals 2025 Sinner - Zverev | * (0.29) | *** (0.72) | ** (0.75) | *** (1.14) |
| | NWKc ATP Finals 2025 Final | n.s. (-0.21) | *** (0.88) | *** (1.51) | ** (0.26) |
| | NWKd ATP Finals 2025 Final | *** (-0.30) | n.s. (0.10) | *** (-0.38) | n.s. (-0.02) |
| | NWKe ATP Finals 2025 Final | * (0.23) | * (0.22) | n.s. (-0.02) | n.s. (-0.25) |
| | NWKg ATP Finals 2025 Final | *** (0.68) | *** (-0.55) | *** (1.59) | * (-0.29) |
| | NWKb Australian Open 2025 Semifinal (Alcaraz - Zverev) | *** (-0.96) | *** (-1.68) | * (-0.18) | * (-0.37) |
| | NWKb Australian Open 2025 Semifinal (Sinner - Djokovic) | n.s. (-0.04) | n.s. (0.11) | n.s. (0.14) | * (0.27) |
| | NWKb Australian Open 2025 Final | *** (0.59) | *** (0.53) | n.s. (-0.09) | * (0.15) |
| | NWKb Serie A 2025/2026 Como 1907 - Atalanta | *** (1.99) | *** (1.54) | *** (1.06) | *** (1.23) |
| | NWKb Serie A 2025/2026 Como 1907 - Fiorentina | *** (-0.58) | *** (0.35) | n.s. (-0.13) | ** (0.29) |

*Continued on next page*

Per-viewing, per-channel comparisons across engagement levels. (continued)

| | Viewing | Ch1 (Lx) VW-CP p-value (Cohen's d) | Ch1 (Lx) VW-BD p-value (Cohen's d) | Ch2 (Rx) VW-CP p-value (Cohen's d) | Ch2 (Rx) VW-BD p-value (Cohen's d) |
|---|---|---|---|---|---|
| | NWKc Australian Open 2025 Semifinal (Sinner - Djokovic) | *** (1.12) | *** (0.81) | *** (1.89) | *** (0.49) |
| | NWKc Australian Open 2025 Semifinal (Alcaraz - Zverev) | *** (1.68) | *** (0.74) | *** (0.64) | * (0.72) |
| | NWKg Serie A 2025/2026 US Cremonese - FC Inter.Milano | *** (1.74) | n.s. (0.06) | *** (-1.75) | * (-0.78) |
| | NWKg Serie A 2025/2026 FC Inter.Milano - Juventus FC | *** (1.38) | *** (0.36) | *** (0.37) | * (0.56) |
| | NWKd Serie A 2025/2026 Juventus FC - SSC Napoli | ** (0.29) | *** (0.49) | ** (-0.18) | *** (0.41) |
| | NWKd Serie A 2025/2026 Parma Calcio 1913 - Juventus FC | n.s. (-0.16) | ** (0.20) | n.s. (0.12) | * (0.82) |
| | NWKd Serie A 2025/2026 FC Inter.Milano - Juventus FC | *** (-0.95) | *** (-0.77) | *** (-0.55) | n.s. (-0.03) |
| | NWKd Australian Open 2025 Semifinal (Sinner - Djokovic) | ** (-0.22) | * (0.18) | *** (0.30) | ** (0.25) |
| @Stadium Viewings | NWKc UEFA Champions League 2025 Semifinal (2nd leg) | *** (-1.25) | *** (0.23) | – – –– | – – –– |
| | NWKc UEFA Champions League 2025 Final | *** (1.19) | *** (-0.54) | – – –– | – – –– |
| | NWKc Serie A 2025/2026 Derby della Madonnina | *** (-2.35) | n.s. (-0.28) | n.s. (0.09) | *** (1.64) |
| | NWKc Serie A 2025/2026 FC Inter.Milano - Bologna FC | *** (-0.24) | n.s. (0.08) | *** (0.29) | *** (0.75) |
| | NWKc Serie A 2025/2026 FC Inter.Milano - Juventus FC | *** (-2.77) | *** (-0.96) | *** (-0.90) | *** (-0.55) |
| Neutral Viewings | NWKd Wimbledon 2025 Final | *** (-0.46) | * (0.14) | *** (-0.55) | n.s. (0.07) |
| | NWKd US Open 2025 Final | * (0.44) | *** (0.68) | *** (-1.21) | n.s. (0.09) |
| | NWKe Wimbledon 2025 Final | *** (-0.66) | n.s. (-0.03) | n.s. (-0.06) | ** (0.37) |
| | NWKe US Open 2025 Final | n.s. (-0.06) | *** (0.36) | *** (-0.76) | *** (-0.83) |

*Continued on next page*

Per-viewing, per-channel comparisons across engagement levels. (continued)

| | Viewing | Ch1 (Lx) VW-CP p-value (Cohen's d) | Ch1 (Lx) VW-BD p-value (Cohen's d) | Ch2 (Rx) VW-CP p-value (Cohen's d) | Ch2 (Rx) VW-BD p-value (Cohen's d) |
|---|---|---|---|---|---|
| | NWKa US Open 2025 Final | n.s. (0.09) | *** (0.65) | *** (-0.37) | *** (0.93) |
| | NWKa ATP Finals 2025 Alcaraz - Musetti | n.s. (0.24) | *** (-0.37) | n.s. (0.30) | n.s. (-0.16) |
| | NWKf US Open 2025 Final | n.s. (0.18) | * (0.23) | n.s. (-0.12) | *** (0.65) |
| | NWKb ATP Finals 2025 Sinner - Zverev | * (0.18) | *** (-0.41) | *** (1.71) | ** (-0.32) |
| | NWKb ATP Finals 2025 Alcaraz - Musetti | *** (0.63) | n.s. (0.04) | *** (2.00) | * (0.38) |
| | NWKb ATP Finals 2025 Sinner - Shelton | *** (-0.74) | *** (0.24) | *** (1.24) | *** (-0.30) |
| | NWKb ATP Finals 2025 Zverev - A.Aliassime | *** (-0.96) | *** (-0.25) | *** (1.75) | n.s. (0.03) |
| | NWKb ATP Finals 2025 Alcaraz - A.Aliassime | *** (-0.66) | *** (-1.05) | *** (2.10) | *** (0.84) |
| | NWKh ATP Finals 2025 Final | *** (-0.25) | *** (-0.61) | – – – – | – – – – |
| | NWKg Australian Open 2025 Semifinal (Alcaraz - Zverev) | *** (0.62) | *** (1.61) | $***(0.52)$ | $***(0.53)$ |
| | NWKd Australian Open 2025 Final | n.s. (-0.13) | n.s. (0.06) | $***(-0.74)$ | $***(-0.81)$ |
| | NWKc Serie A 2025/2026 US Cremonese - FC Inter.Milano | *** (-1.80) | *** (0.70) | $***(0.37)$ | $**(-0.30)$ |
| | NWKc Australian Open 2025 Final | *** (-3.58) | *** (-0.56) | *n.s.*(0.22) | *n.s.*(0.13) |
| Non-Viewings | NWKd UEFA Champions League 2025 Semifinal (2nd leg) | n.s. (-0.07) | n.s. (-0.14) | – – – – | – – – – |
| Non-Viewings | NWKd UEFA Champions League 2025 Final | n.s. (0.27) | n.s. (0.07) | – – – – | – – – – |
| Non-Viewings | NWKd Roland Garros 2025 Final | *** (-0.29) | * (-0.03) | – – – – | – – – – |
| Non-Viewings | NWKe UEFA Champions League 2025 Semifinal (2nd leg) | *** (-0.29) | *** (-0.62) | *** (0.85) | *** (0.78) |
| Non-Viewings | NWKe UEFA Champions League 2025 Final | *** (0.40) | *** (0.58) | n.s. (0.06) | n.s. (0.46) |
| Non-Viewings | NWKe Roland Garros 2025 Final | *** (0.62) | *** (1.00) | *** (0.23) | *** (0.72) |

*Continued on next page*

Per-viewing, per-channel comparisons across engagement levels. (continued)

| **Viewing** | **Ch1 (Lx)** VW-CP p-value (Cohen's d) | VW-BD p-value (Cohen's d) | **Ch2 (Rx)** VW-CP p-value (Cohen's d) | VW-BD p-value (Cohen's d) |
|---|---|---|---|---|
| NWKf<br>UEFA Champions League<br>2025 Semifinal (2nd leg) | *** (-0.50) | n.s. (0.09) | – – –– | – – –– |
| NWKf Wimbledon<br>2025 Final | *** (-0.32) | *** (-0.43) | *** (-0.39) | *** (-0.42) |
| NWKb<br>UEFA Champions League<br>2025 Final | n.s. (-0.18) | * (-0.24) | *** (-1.39) | *** (-0.75) |
| NWKc Roland Garros<br>2025 Final | *** (0.49) | *** (-0.91) | – – –– | – – –– |
| NWKa Roland Garros<br>2025 Final | *** (-0.41) | *** (0.55) | *** (-0.29) | n.s. (-0.22) |
| NWKa Wimbledon<br>2025 Final | n.s. (0.12) | *** (-0.85) | n.s. (0.01) | n.s. (0.18) |

| | | | | | |
|---|---|---|---|---|---|
| p-value | $> 0.5$<br>(n.s.) | $< 0.005$<br>(***) | $< 0.01$<br>(**) | $< 0.05$<br>(*) | no data<br>(– – ––) |
| Cohen's d | ($-0.10 < d < 0.10$ or n.s.)<br>Negligible | $d \geq 0.7$<br>$d \leq -0.7$<br>Large | $d \geq 0.3$<br>$d \leq -0.3$<br>Medium | $d \geq 0.1$<br>$d \leq -0.1$<br>Small | (– – ––)<br>no data |

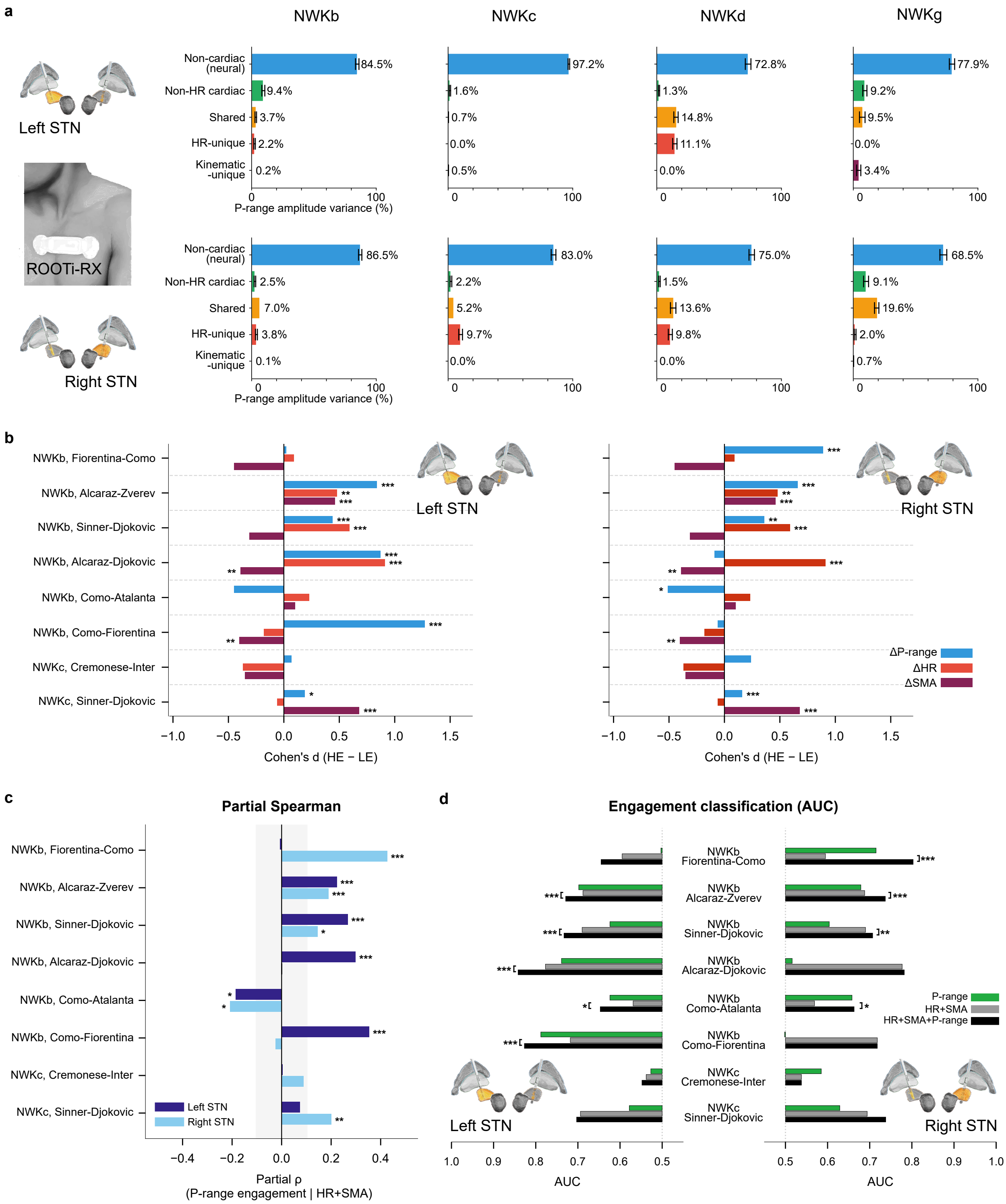


**Fig. 8**: **P-range neural signal variance decomposition and engagement non-redundancy analyses. a,** P-range amplitude variance decomposed into five components via OLS regression for left and right STN of four patients for whom ROOTi-RX recordings were available (NWKb, NWKc, NWKd, NWKg) across the full concurrent cardiac and kinematic monitoring period. Components include: non-cardiac (neural) variance unexplained by any physiological covariate; non-HR cardiac variance attributable to autonomic indices (RMSSD, SDNN, LF/HF, EDR) beyond heart rate; shared variance between HR and other cardiac indices; HR-unique variance providing an upper bound on rate-dependent cardiac artifact; kinematic-unique variance attributable to chest-mounted accelerometer (signal magnitude area, SMA). Error bars denote 95% bootstrap confidence intervals. **b,** Cohen's d (difference between high-engagement HE and low-engagement LE intervals) for P-range amplitude (ΔP-range), heart rate (ΔHR), and SMA (ΔSMA) across eight viewings (six episodes for NWKb, two episodes for NWKc), shown separately for left and right STN. Asterisks denote significance ($^{*}p < 0.05$, $^{**}p < 0.01$, $^{***}p < 0.005$, Mann-Whitney U test). *(continues...)*

**Fig. 8**: *(...continues)* **c,** Partial Spearmann correlation between P-range amplitude and engagement after controlling for HR and SMA, for left and right STN. **d,** Logistic regression AUC for engagement classification using P-range amplitude alone, HR+SMA, and HR+SMA+P-range, shown separately by hemisphere. Bars extending leftward from 0.5 (left STN) and rightward from 0.5 (right STN) indicate performance above chance. Asterisks on brackets denote significant AUC improvement when P-range is added to the physiological model (likelihood-ratio test, dof= 1).

**Table Supplementary Information 4**:
Refinement of HR-unique neural variance component from OLS decomposition.

| Patients | n (min) | STN Hem. | HR → P-range F-statistic (p-value) | P-range → HR F-statistic (p-value) | HR-unique% day/night | SMA → P-range F-statistic (p-value) | P-range → SMA F-statistic (p-value) | Kinematic-unique% day/night |
|---|---|---|---|---|---|---|---|---|
| NWKb | 7835 | L | Bidirectional | | 1.56 / 0.62 | Bidirectional | | 0.02 / 0.07 |
| | | | 33.70 (lag=4) (***) | 14.76 (lag=1) (***) | | 3.70 (lag=4) (**) | 3.26 (lag=9) (***) | |
| | | R | HR → P-range | | 2.52 / 4.47 | None | | 0.08 / 0.17 |
| | | | 45.11 (lag=1) (***) | 1.78 (lag=2) (n.s.) | | 2.12 (lag=4) (n.s.) | 1.57 (lag=9) (n.s.) | |
| NWKc | 6654 | L | Bidirectional | | 0.91 / 2.43 | P-range → SMA | | 1.18 / 2.97 |
| | | | 10.03 (lag=3) (***) | 16.71 (lag=1) (***) | | 0.63 (lag=1) (n.s.) | 5.08 (lag=1) (*) | |
| | | R | Bidirectional | | 9.17 / 6.61 | None | | 0.03 / 0.11 |
| | | | 15.73 (lag=1) (***) | 8.93 (lag=2) (***) | | 0.45 (lag=1) (n.s.) | 1.35 (lag=10) (n.s.) | |
| NWKd | 6585 | L | P-range → HR | | 5.51 / 18.10 | Bidirectional | | 0.02 / 0.02 |
| | | | 0.90 (lag=6) (n.s.) | 17.78 (lag=6) (***) | | 3.58 (lag=2) (*) | 3.81 (lag=1) (*) | |
| | | R | Bidirectional | | 8.39 / 12.89 | SMA → P-range | | 0.03 / 0.11 |
| | | | 6.39 (lag=1) (*) | 50.70 (lag=3) (***) | | 3.94 (lag=2) (*) | 1.50 (lag=8) (n.s.) | |
| NWKg | 3483 | L | P-range → HR | | 5.52 / 3.84 | None | | 0.35 / 0.02 |
| | | | 1.23 (lag=1) (n.s.) | 2.42 (lag=8) (*) | | 0.28 (lag=1) (n.s.) | 1.04 (lag=7) (n.s.) | |
| | | R | Bidirectional | | 0.58 / 1.34 | None | | 0.13 / 0.60 |
| | | | 69.55 (lag=1) (***) | 2.06 (lag=9) (*) | | 0.00 (lag=1) (n.s.) | 1.70 (lag=7) (n.s.) | |

(*) p-value $< 0.05$; (**) p-value $< 0.01$; (***) p-value $< 0.005$; (n.s.) not significant; .→. Granger causality direction.

**Table Supplementary Information 5**:
Neural and cardiac comparison of within-match Higher vs Low Engagement intervals (HE vs LE).

| **Patients** | **Viewing** | **HE/LE (min)** | **STN Hem.** | **d($\Delta P$-range) [p-value]** | **d($\Delta\|dP$-range$/dt\|$) [p-value]** | **d($\Delta HR$) [p-value]** | **d($\Delta SMA$) [p-value]** |
|---|---|---|---|---|---|---|---|
| NWKb | Coppa Italia 25/26<br>Fiorentina-Como | 35/73 | L<br>R | +0.02 [n.s.]<br>+0.89 [***] | -0.06 [n.s.]<br>-0.49 [*] | +0.09 [n.s.] | -0.45 [n.s.] |
| | Serie A 25/26<br>Como-Atalanta | 25/93 | L<br>R | -0.45 [n.s.]<br>-0.51 [*] | +1.01 [***]<br>+0.26 [n.s.] | +0.23 [n.s.] | +0.10 [n.s.] |
| | Australian Open 25/26<br>(Semif.) Alcaraz-Zverev | 85/244 | L<br>R | +0.84 [***]<br>+0.66 [***] | +0.32 [**]<br>+0.23 [n.s.] | +0.48 [**] | +0.46 [***] |
| | Australian Open 25/26<br>(Semif.) Sinner-Djokovic | 129/121 | L<br>R | +0.44 [***]<br>+0.36 [**] | +0.42 [***]<br>+0.21 [n.s.] | +0.59 [***] | -0.31 [n.s.] |
| | Australian Open 25/26<br>(Final) Alcaraz-Djokovic | 44/143 | L<br>R | +0.87 [***]<br>-0.09 [n.s.] | +0.85 [***]<br>+0.04 [n.s.] | +0.91 [***] | -0.39 [**] |
| | Serie A 25/26<br>Como-Fiorentina | 16/97 | L<br>R | +1.27 [***]<br>-0.06 [n.s.] | +0.64 [**]<br>+0.97 [**] | -0.18 [n.s.] | -0.40 [**] |
| NWKc | Australian Open 25/26<br>(Semif.) Sinner-Djokovic | 112/138 | L<br>R | +0.19 [*]<br>+0.16 [***] | +0.34 [**]<br>+0.21 [n.s.] | -0.06 [n.s.] | +0.68 [***] |
| | Serie A 25/26<br>Cremonese-Inter | 32/84 | L<br>R | +0.07 [n.s.]<br>+0.24 [n.s.] | +0.22 [n.s.]<br>-0.18 [n.s.] | -0.37 [n.s.] | -0.35 [n.s.] |

| **Cross-viewing analysis n=8** | **STN Hem.** | $\Delta\|dP$**-range**$/dt\|$ **vs** $\Delta HR$ $\rho$ **[p-value]** | **d($\Delta P$-range) vs d($\Delta HR$)** $\rho$ **[p-value]** | **d($\Delta P$-range) vs d($\Delta SMA$)** $\rho$ **[p-value]** |
|---|---|---|---|---|
| | L | +0.25 [n.s.] | -0.71 [*] | +0.00 [n.s.] |
| | R | +0.41 [n.s.] | -0.29 [n.s.] | +0.05 [n.s.] |

[*] p-value $<$ 0.05; [**] p-value $<$ 0.01; [***] p-value $<$ 0.005; [n.s.] not significant; d(.) Cohen's d.
